# Combined genome-wide association studies in humans and cattle: Cross-species genetic insights into dizygotic twinning

**DOI:** 10.64898/2026.11.16.26367253

**Authors:** Nikki Hubers, Marijn Schipper, Scott Gordon, Jaakko Leinonen, Árni Gunnarsson, Christian M Page, Marianne Nygaard, Charlotte KL Pahnke, Robert Karlsson, Teemu Palviainen, Jamee M Berg, Tanguy Rubat du Mérac, Kaito Kawakami, Wangjingyi Liao, Elham Assary, Maryam Kazemi Naeini, Martje Bos, Ilja M Nolte, Peter J van der Most, Lifelines Cohort Study, Scott Gallagher, Samantha LP Schilit, Anastasia Goula, Yuri Milaneschi, René Pool, Jenny van Dongen, Christiaan de Leeuw, Jeffrey Beck, Erik Ehli, Vera Fuentes-Moreno, Emma Vlasuik, Yoon-Mi Hur, Bruno Reversade, Velja Mijatovic, Claudio D Stern, Harold Snieder, Kaare Christensen, Frances Williams, Robert Plomin, Miina Ollikainen, Patrik KE Magnusson, Mirko Ruks, Carlo Maj, Christian Kandler, Andreas J. Forstner, Cynthia C Morton, Daniel I Chasman, William Iacono, Jennifer R Harris, Taru Tukianen, Martin Kennedy, Gonneke Willemsen, Cornelis B Lambalk, Hreinn Stefansson, Jaakko Kaprio, Grant W Montgomery, Jouke-Jan Hottenga, Hamdi Mbarek, Christin Schmidtmann, Laura Hüneke, Donnagh P. Berry, Brian W Kirkpatrick, Nicholas G Martin, Dorret I Boomsma

**Author notes:** These authors contributed equaly (Nikki Hubers, Marijn Schipper). Author information in Supplementary data 1 (Lifelines Cohort Study).

## Abstract

To leverage independent genetic evidence from multiple species, we developed a cross-species gene-based GWAS meta-analysis framework and applied this to Dizygotic Twinning (DZT) in humans and cattle. After meta-analysis within species, we converted human and cattle GWAS summary statistics to gene-level association signals within each species, mapped genes across species through human–cattle orthology, and then combined gene-level evidence for orthologous genes across the two species. DZT is defined as giving birth to multiple offspring after a single pregnancy resulting from spontaneous multiple ovulation. In humans, DZT occurs in about 1% of pregnancies with heritability estimates of 20% to 30% whilst in cattle DZT occurs in 1-5% of pregnancies and heritability is estimated at about 11%. Here, we present the largest human GWAS meta-analysis including data from 1,395,285 participants. We identified 16 genetic regions of interest, including 12 independent loci from the SNP based analyses and 4 genes identified in the gene-based MAGMA analyses. Among the prioritized genes are follicle stimulation hormone receptor (FSHR) and growth differentiation factor 9 (GDF9), that both are involved in ovarian folliculogenesis. Cattle GWAS summary statistics were based on data from 2,656 Irish Holstein bulls with estimated breeding values for predisposition to twinning based on 2,238,936 birth records, and 86,391 German Holstein dams. After combining the gene-level analyses for 13,225 genes mapped as orthologous across the human and cattle data we identify FSHR as a central locus for DZT in both humans and cattle. We find evidence of additional associations, including ACVR2A, which plays a role in embryonic implantation and TUFM, which has previously been implicated in premature ovarian insufficiency. Our approach provides a promising framework for future cross-species comparative GWAS studies. It shows benefits for gene discovery and gene prioritization and in our application expands available functional follow-up data, allowing us to leverage cattle eQTL data to resolve human GWAS loci. The method and underlying philosophy presented here are transferable to other shared phenotypes across several species.

## Introduction

Dizygotic twinning (DZT) is defined as giving birth to multiple offspring after a single pregnancy resulting from a spontaneous multiple ovulation.^1,2^ DZT has been observed as a heritable trait in both humans and other mono-ovulatory mammals.^3–6^ Currently, well-powered genome-wide association studies (GWAS) of DZT exist for humans and cattle.^2,7,8^ We therefore decided to explore to what extent the genetic regulation of DZT is shared across species, and whether a combined meta-analysis could help solve some of the challenges in interpreting the results from GWAS. To this end, we perform the largest human GWAS for DZT to date and explore the feasibility of cross-species meta-analysis by combining human and cattle data.

The trait of multiple ovulation is regulated through complex interactions among endocrine signals and intra-ovarian signaling pathways that govern follicular development. Key components of these regulatory networks were identified through early endocrine studies, mouse knockout models, and genetic analyses in sheep, cattle and humans.^5,6,9^ Central to this process are variations in circulating follicle-stimulating hormone (FSH) concentrations during follicular selection, oocyte-derived signaling mediated by growth differentiation factor 9 (GDF9) and bone morphogenetic protein 15 (BMP15), and the temporal dynamics of FSH receptor (FSHR) and luteinizing hormone receptor (LHCGR) expression during follicular maturation.^6,9^ Collectively, these mechanisms determine the number of follicles that survive the selection process and proceed to ovulation.

GWASs into human DZT provided support for the central role of hypothalamic-pituitary-ovarian signaling in the regulation of DZT and ovulation.^2,10^ Our 2024 GWAS including mothers of DZ twins and DZ twins as cases identified four loci associated with DZT: *FSHB*, *SMAD3*, *GNRH*, and *ZMFP1.*^2,11^ Gene-based testing and functional follow-up also indicated roles for *FSHR* and *IPO8*. These genes are involved in the follicle-stimulating hormone (FSH) and transforming growth factor beta (*TGF-β*) pathways, which regulate circulating FSH levels, sensitivity to FSH and ovulation.^12,13^ The SNP-based heritability (h^2^) was estimated at 2.4%,^2^ while heritability estimates from pedigree studies for human DZT range between 20-30%.^14^

In Holstein dairy cattle, the SNP-based heritability estimates for multiple births are similar to those in humans (1.9-2.6%).^7^ There is a specific interest in DZT in dairy cattle as DZT is economically disadvantageous for farmers.^15^ For example, in opposite-sex twin offspring, freemartinism as characterized by underdeveloped reproductive organs and masculinized features due to in-utero hormonal exchange between the twins, often results in infertile female offspring.^16^ A previous GWAS identified a locus on bovine chromosome 11 containing the FSH receptor (*FSHR*) and Luteinizing Hormone/Choriogonadotropin Receptor (*LHCGR*) as associated with DZT in an Irish sample of 2,656 Holstein cattle.^7,17^

These parallels—spanning both genetic architecture and highly similar phenotypes across humans and cattle—motivated a cross-species GWAS meta-analysis. Meta-analysis typically relies on overlapping SNP markers, which are rare across species. For instance between chimpanzees and humans, which share a relatively recent common ancestor, only ∼0.2–0.3% of variants are shared,^18^ and the overlap between humans and cattle can be expected to be smaller still. The number of conserved orthologous genes, i.e. conserved genes shared with species inherited from a common ancestor is much larger, with about 72-76% of genes having a direct one-to-one ortholog.^19^ The extensive conservation of orthologous genes across these species opens the possibility of gene-level meta-analysis as a viable and powerful alternative.

Earlier studies have investigated the overlap between gene sets associated with fertility and body size in humans, dogs and cattle.^20–23^ These were not gene discovery designs, but rather focused on cross-species overlap functions as candidate-gene prioritization and pathway-convergence tools, clearly establishing that multi-species GWAS comparisons can help identify conserved growth mechanisms underlying stature/height and fertility. For instance, an 87-gene set from cattle was tested for cross-species convergence with results from a human reproductive-timing GWAS, identifying significant enrichment of shared genes.^20^ Eight of the cattle fertility-related genes that were associated with age at natural menopause and four with age at menarche. This overlap exceeded chance expectations and supports a shared pool of genes influencing fertility-related biology across mammals.

To our knowledge, no formal full cross-species meta-analyses of GWAS data have been conducted previously. Integrating GWAS results from humans and cattle has the potential to enhance genetic discovery in both species. Human populations generally exhibit shorter linkage disequilibrium (LD) blocks than commercial cattle populations, where extensive LD can hinder fine-mapping efforts.^24^ In addition, combining evidence across species is expected to increase statistical power to detect loci involved in conserved biological pathways. Cross-species analyses may also facilitate downstream functional interpretation. Extensive expression quantitative trait locus (eQTL) resources are available for a diverse range of tissues in cattle and other livestock species, including tissues that are difficult or impossible to obtain from living humans.^25^ Access to reproductive tissues and experimental populations in cattle enables functional validation of candidate genes and regulatory mechanisms.^17^ Therefore, we propose a cross-species gene-level GWAS framework that could leverage complementary genomic and transcriptomic resources across species to accelerate the identification and characterization of genes and apply this framework to DZT and ovulation rate.

In our study we take a stepwise approach by first presenting the GWAS meta-analysis in humans and secondly perform cross-species genome-wide gene-based association tests (using MAGMA)^26^ based on the newly presented GWAS results of DZT in humans and two cattle GWASs. In the human GWAS we increased discovery sample to 1,395,285 participants from European ancestry (Table 1), which included 13,554 mothers of naturally conceived DZ twin cases, 41,550 DZ twin cases and 1,340,181 controls, leading to an effective sample size of 131,893 (Methods). We include data from 10 different countries and 15 separate cohorts. The two cattle GWASs were performed in 2,656 Irish Holstein bulls with estimates of genetic merit for twinning from 2,238,936 calving records, and a cohort of 86,391 German Holstein dams with twinning records.^7^ We meta-analyze the cattle MAGMA Z-scores using a Stouffer’s Z, weighted based on attenuation ratio, and subsequently meta-analyze the cattle and human MAGMA Z-scores via an unweighted Stouffer’s Z, restricting to genes that are orthologous between the two species.^26–28^

**Table 1:** Overview of human samples in the Genome-wide association meta-analysis.

| **Mothers of DZ twins** | **Cases** | **Controls** | **Total** |
|---|---|---|---|
| Netherlands Twin Register and Prospect (Netherlands) | 1808 | 6373 | 8181 |
| QIMR (Australia) | 4942 | 29327 | 34269 |
| MCTFR (United States) | 568 | 1862 | 2430 |
| WGHS (United States) | 361 | 20183 | 20544 |
| deCODE (Iceland) | 2877 | 212767 | 215644 |
| FinnGen (Finland) | 2009 | 478349 | 480358 |
| Moba (Norway) | 658 | 53046 | 53704 |
| TwinLife (Germany) | 331 | 660 | 991 |
| **Total MoDZTs** | **13554** | **802567** | **816121** |
| **DZ twin individuals** | **Cases** | **Controls** | **Total** |
| UK Biobank (United Kingdom) | 8962 | 409591 | 418553 |
| Swedish Twin Registry | 15251 | 3420 | 18671 |
| Finnish Twins Cohort | 6021 | 2167 | 8188 |
| TEDS (United Kingdom) | 2919 | 2580 | 5499 |
| Twins UK (United Kingdom) | 2206 | 3491 | 5697 |
| Danish Twin Registry | 1189 | 391 | 1580 |
| Netherlands Twin Register | 3810 | 7695 | 11505 |
| Lifelines (Netherlands) | 718 | 67657 | 68375 |
| MoBa (Norway) | 474 | 40622 | 41096 |
| **Total DZ twins** | **41550** | **537614** | **579164** |
| ***Total DZ twins + MoDZTs*** | ***55104*** | ***1340181*** | ***1395285*** |

## Results

### Updated meta-analysis of human DZT identifies novel DZ twinning loci

We performed a GWAS meta-analysis of 15 human cohorts with DZT data, based on 1,395,285 participants from 10 countries (Table 1). The effective sample size was estimated at 131,893 (Methods). In total, we identified 12 independent genetic loci that reach genome-wide significance (Table 2; Fig. 1; Supplementary Fig. 1). Our results include the four previously identified loci, linked to *FSHB*, *SMAD3*, *GNRH*, and *ZFPM1.*^2^ We performed gene-based testing using MAGMA (Supplementary Table 1; Supplementary Fig. 1),^26^ which highlighted four other genes in additional genetic regions. Two of these genes, *IPO8* (z = 5.09; p = 1.80×10^−7^) and *SHBG* (z = 4.69; p= 1.36×10^−6^) have been identified for DZT before and two of these genes, *REXO1* (z = 4.65; p = 1.67x10^-6^) and *SGMS2* (z = 4.72; p = 1.16x10^-6^) are new findings. An overview of the 16 genetic regions, with the 12 genome-wide significant loci and the 4 additional MAGMA significant genes, and the most likely causal gene within this region, is provided in Table 3.

**Fig. 1:**
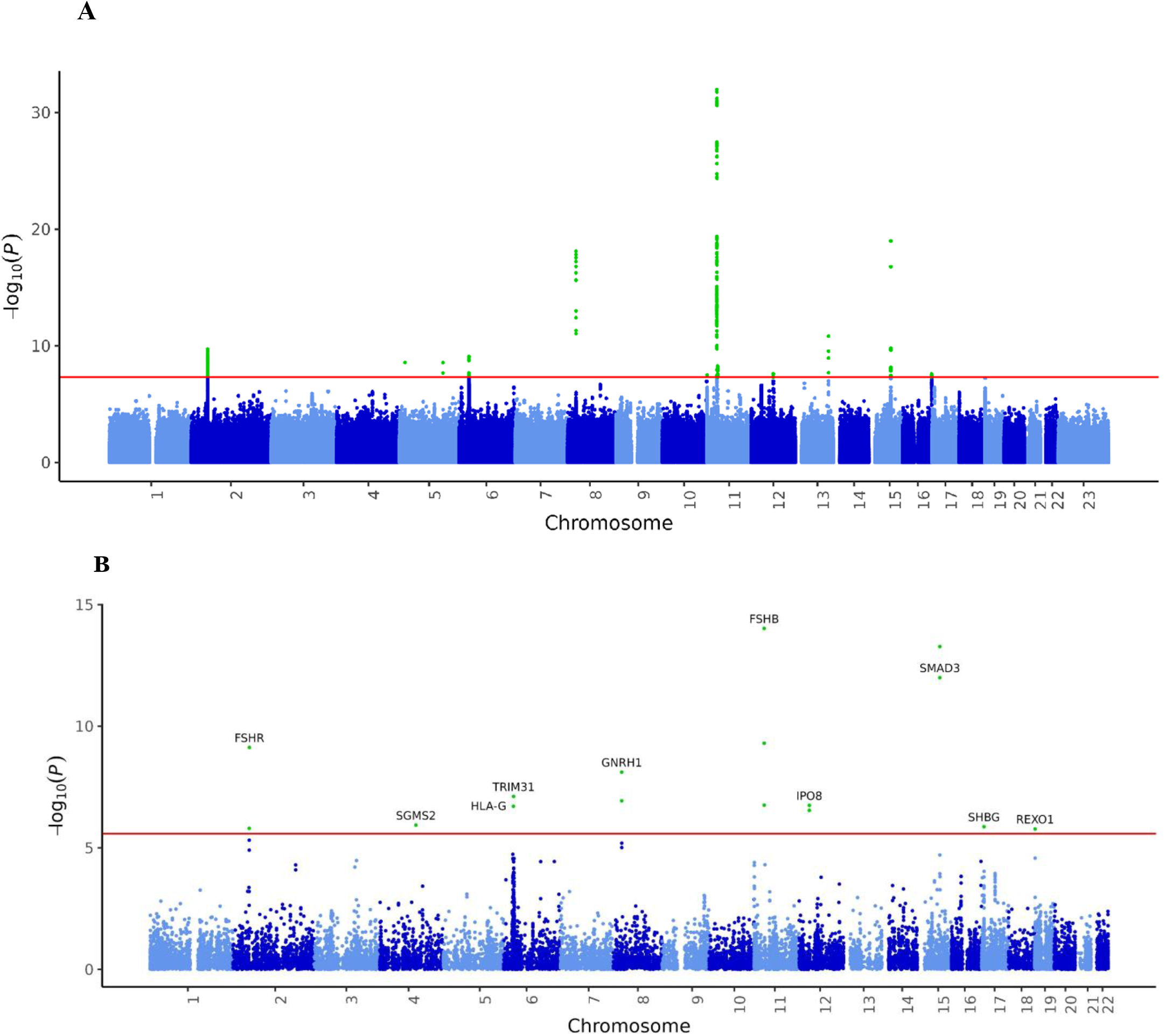
Association results of the genome-wide association study (GWAS) meta-analysis of human dizygotic (DZ) twinning versus controls. **A**) Manhattan plot of single-nucleotide polymorphisms (SNP) associations. The dashed red line represents the genome-wide level of significance (P value< 5 × 10^−8^). **B**) Manhattan plot of gene-based association results as computed by MAGMA. The -log10P value (y-axis) for the genes is plotted against their physical chromosomal position (x-axis). Input SNPs were mapped to 19,171 protein-coding genes. Genome-wide significance (red dashed line in the plot) was defined at P = 0.05/19,171 = 2.61 × 10^−6^.

**Table 2:**
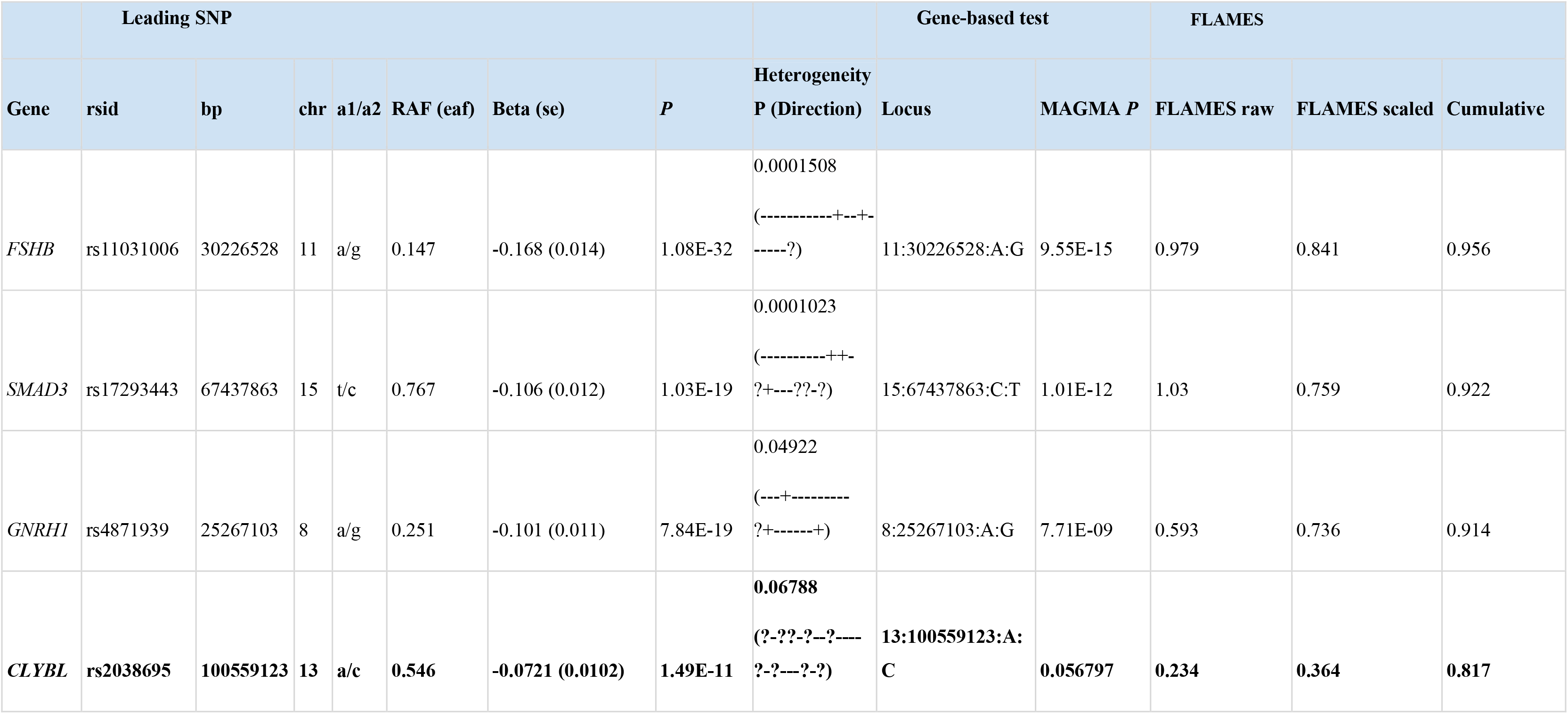
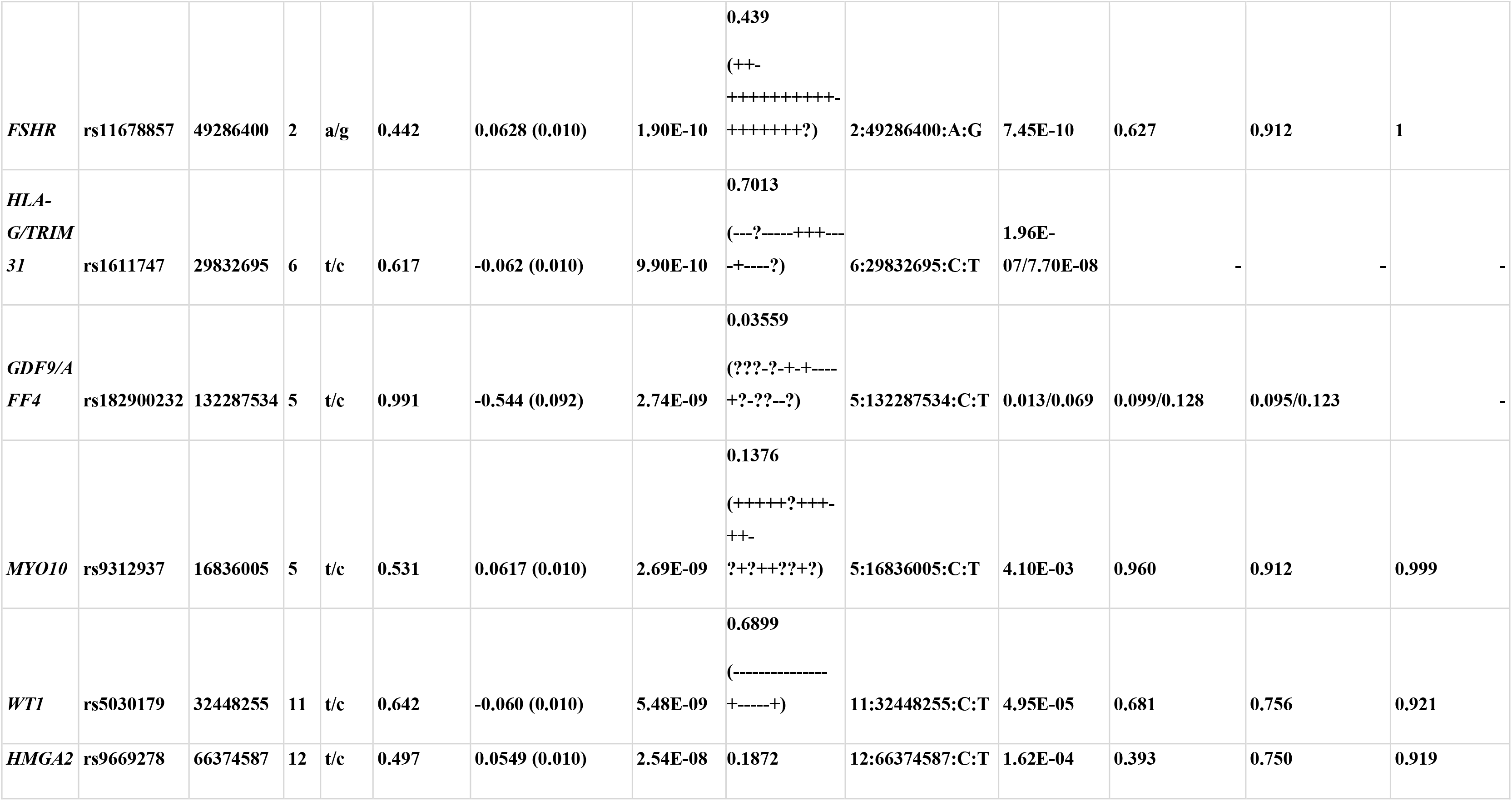
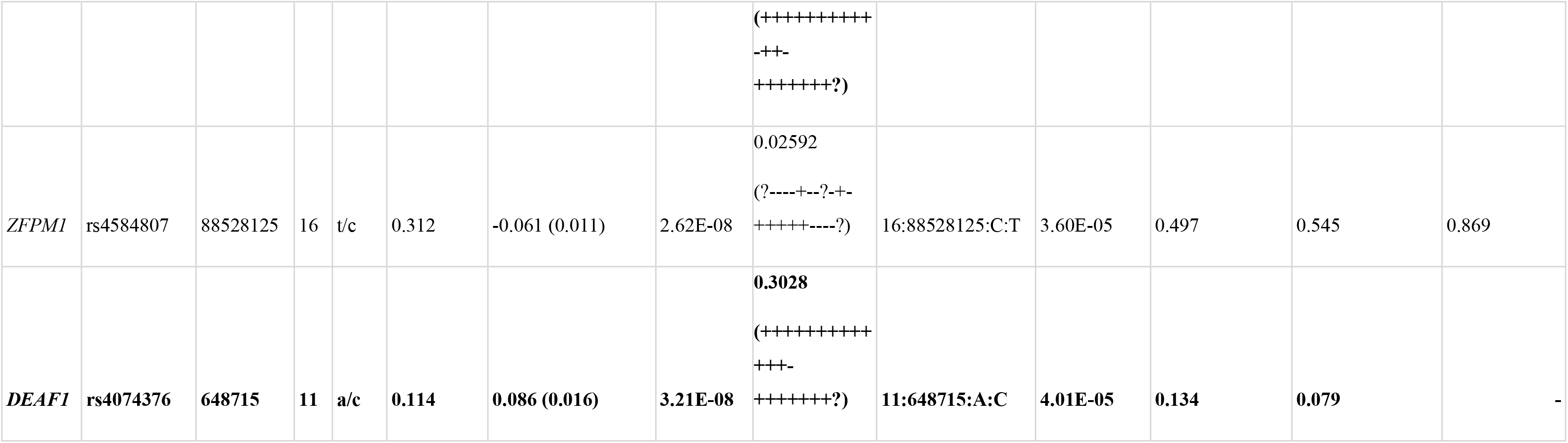
Overall human genome-wide association meta-analysis results and the MAGMA and FLAMES results from the significant loci. Bp; base pair, chr; chromosome, A1: Effect allele as used in the statistical analyses: A2: Other allele EAF; effect allele frequency. The locus column represents chromosome: base pair: reference allele: other allele. The Heterogeneity indicates the P value from the Cochran’s Q-test. In **bold** are the SNPs that were not implicated in the GWAS meta-analyses by Mbarek et al., 2024.^2^

|  | **Leading SNP** |  |  |  |  |  |  |  | **Gene-based test** |  | **FLAMES** |  |  |
|---|---|---|---|---|---|---|---|---|---|---|---|---|---|
| **Gene** | **rsid** | **bp** | **chr** | **a1/a2** | **RAF (eaf)** | **Beta (se)** | ***P*** | **Heterogeneity P (Direction)** | **Locus** | **MAGMA *P*** | **FLAMES raw** | **FLAMES scaled** | **Cumulative** |
| *FSHB* | rs11031006 | 30226528 | 11 | a/g | 0.147 | -0.168 (0.014) | 1.08E-32 | 0.0001508<br>(-----------+--+------?) | 11:30226528:A:G | 9.55E-15 | 0.979 | 0.841 | 0.956 |
| *SMAD3* | rs17293443 | 67437863 | 15 | t/c | 0.767 | -0.106 (0.012) | 1.03E-19 | 0.0001023<br>(----------++-?+---??-?) | 15:67437863:C:T | 1.01E-12 | 1.03 | 0.759 | 0.922 |
| *GNRH1* | rs4871939 | 25267103 | 8 | a/g | 0.251 | -0.101 (0.011) | 7.84E-19 | 0.04922<br>(---+---------?+------+) | 8:25267103:A:G | 7.71E-09 | 0.593 | 0.736 | 0.914 |
| ***CLYBL*** | **rs2038695** | **100559123** | **13** | **a/c** | **0.546** | **-0.0721 (0.0102)** | **1.49E-11** | **0.06788<br>(?-??-?--?----?-?---?-?)** | **13:100559123:A:C** | **0.056797** | **0.234** | **0.364** | **0.817** |
| ***FSHR*** | **rs11678857** | **49286400** | **2** | **a/g** | **0.442** | **0.0628 (0.010)** | **1.90E-10** | **0.439<br>(++-++++++++++-+++++++?)** | **2:49286400:A:G** | **7.45E-10** | **0.627** | **0.912** | **1** |
| ***HLA-G/TRIM31*** | **rs1611747** | **29832695** | **6** | **t/c** | **0.617** | **-0.062 (0.010)** | **9.90E-10** | **0.7013<br>(---?-----+++----+----?)** | **6:29832695:C:T** | **1.96E-07/7.70E-08** | **-** | **-** | **-** |
| ***GDF9/AFF4*** | **rs182900232** | **132287534** | **5** | **t/c** | **0.991** | **-0.544 (0.092)** | **2.74E-09** | **0.03559<br>(???-?-+-+----+?-??--?)** | **5:132287534:C:T** | **0.013/0.069** | **0.099/0.128** | **0.095/0.123** | **-** |
| ***MYO10*** | **rs9312937** | **16836005** | **5** | **t/c** | **0.531** | **0.0617 (0.010)** | **2.69E-09** | **0.1376<br>(+++++?+++-++-?+?++??+?)** | **5:16836005:C:T** | **4.10E-03** | **0.960** | **0.912** | **0.999** |
| ***WT1*** | **rs5030179** | **32448255** | **11** | **t/c** | **0.642** | **-0.060 (0.010)** | **5.48E-09** | **0.6899<br>(--------------+-----+)** | **11:32448255:C:T** | **4.95E-05** | **0.681** | **0.756** | **0.921** |
| ***HMGA2*** | **rs9669278** | **66374587** | **12** | **t/c** | **0.497** | **0.0549 (0.010)** | **2.54E-08** | **0.1872<br>(++++++++++-++-+++++++?)** | **12:66374587:C:T** | **1.62E-04** | **0.393** | **0.750** | **0.919** |
| *ZFPM1* | rs4584807 | 88528125 | 16 | t/c | 0.312 | -0.061 (0.011) | 2.62E-08 | 0.02592<br>(?----+--?-+-+++++----?) | 16:88528125:C:T | 3.60E-05 | 0.497 | 0.545 | 0.869 |
| ***DEAF1*** | **rs4074376** | **648715** | **11** | **a/c** | **0.114** | **0.086 (0.016)** | **3.21E-08** | **0.3028<br>(+++++++++++++-+++++++?)** | **11:648715:A:C** | **4.01E-05** | **0.134** | **0.079** | **-** |

**Table 3:** Gene names, and previous associations based on the GWAS catalogue, of the 16 genetic regions of interest identified in the human DZT GWAS. The 16 regions include the 12 independent loci from the SNP based analyses with the prioritized genes from the FLAMES analyses and the 4 genes identified in the gene-based MAGMA analyses.

| **Causal gene (FLAMES or MAGMA)** | **Human Chr (chr cattle)** | **Gene Name** | **Previous associations relevant traits** | **P Magma Humans** | **P Magma cattle** | **P Cross-species MA** |
|---|---|---|---|---|---|---|
| *FSHR* | 2 (11) | Follicle stimulating hormone receptor | FSH levels, PCOS/PMOS, Height<sup>108–110</sup> | 7.45E-10 | 5.62e-19 | 3.77e-26 |
| *SGMS2* | 4 (6) | Sphingomyelin Synthase 2 | Height, Breastfeeding duration<sup>110,111</sup> | 1.16E-06 | 0.2809528 | 8.83e-05 |
| *MYO10* | 5 (20) | Unconventional Myosin-10 | Age at menarche, PCOS/PMOS, Height<sup>112,113</sup> | 4.10E-03 | 0.9286235 | 0.2023737 |
| *GDF9* | 5 (5) | Growth differentiation factor 9 | Height<sup>110</sup> | 0.01311 | 4.23e-05 | 6.77e-06 |
| *HLA-G* | 6 (23) | Major histocompatibility complex, class I, G | Waist-to-hip ratio adjusted for BMI<sup>114</sup> | 1.96E-07 | - | - |
| *GNRH1* | 8 (14) | Gonadotropin releasing hormone 1 | Age at Menopause<sup>115</sup> | 7.71E-09 | 0.2643871 | 4.38883e-06 |
| *FSHB* | 11 (15) | Follicle stimulating hormone subunit-b | FSH levels, PCOS/PMOS, Endometriosis<sup>54–56</sup> | 9.55E-15 | 0.8320248 | 1.10255e-06 |
| *WT1* | 11 (15) | Wilms' Tumor factor 1 | Waist-hip ratio, Endometriosis, Ovarian cyst, Height, Age at menopause<sup>110,112,115–117</sup> | 4.95E-05 | 0.06108978 | 6.01e-05 |
| *DEAF1* | 11 (29) | Deformed epidermal autoregulatory factor-1 | Height<sup>110,112</sup> | 4.01E-05 | 0.5870848 | 0.00422799 |
| *HMGA2* | 12 (5) | High mobility group AT-hook 2 | Height, SHBG levels, Smoking cessation, Waist-to-hip ratio adjusted for BMI<sup>112,118–120</sup> | 0.000162 | - | - |
| *IPO8* | 12 (5) | Importin 8 | Waist-to-hip ratio adjusted for BMI, Height, Breastfeeding duration<sup>111,112,121</sup> | 1.80E-07 | 0.8441915 | 0.001968709 |
| *CLYBL* | 13 (12) | Citramalyl-CoA lyase | Smoking (ever), Testosterone levels, BMI, Height, Age at Menopause<sup>110,112,118,122,123</sup> | 0.056797 | 0.3897396 | 0.09395743 |
| *SMAD3* | 15 (10) | SMAD family member 3 | Height, Waist-to-hip ratio adjusted for BMI, Smoking initiation<sup>110,112,119</sup> | 1.01E-12 | 0.5415061 | 4.812143e-07 |
| *ZFPM1* | 16 (18) | Zinc Finger Protein, FOG Family Member 1 | SHBG levels, FSH levels, Age at Menopause<sup>112,118,124</sup> | 3.60E-05 | 0.2840933 | 0.0006625351 |
| *SHBG* | 17 (19) | Sex hormone binding globulin | Testosterone levels, SHBG levels, Estrogen levels, Height, Waist-hip ratio, Waist-to-hip ratio adjusted for BMI, Smoking cessation<sup>110,112,118,119,125,126</sup> | 1.36E-06 | 0.7934544 | 0.003085277 |
| *REXO1* | 19 (7) | RNA Exonuclease 1 Homolog | Testosterone levels, BMI, Age at Menarche<sup>112,118,122</sup> | 1.67E-06 | 0.01155304 | 4.959651e-07 |

For each of the 12 significant loci in the human DZT GWAS, we performed gene prioritization with the FLAMES software.^29^ For each locus, FLAMES annotates fine-mapped credible sets and applies a machine learning classifier to score each gene, where this score denotes the level of biological evidence for that gene being regulated by a set of credible causal SNPs in the locus. For most loci, FLAMES was able to predict a likely causal gene (Table 2; Supplementary Table 2) above the confidence threshold, except for the rs182900232 locus on chromosome 5, rs1611747/rs1362104 locus on chromosome 6 and rs4074376 on chromosome 11. The locus zoom plots and the FLAMES decision trees for these three loci are provided in Supplementary Fig. 2-4. Rs182900232 on chromosome 5 is located within the *AFF4* gene, even though as explained below, the nearby *GDF9* gene is a more likely candidate based on previous work (*Fine-mapping in the Cross-species analysis*).^30,31^ The gene-based tests indicate that the most likely candidates for the rs1611747/rs1362104 locus on chromosome 6 are *HLA-G* and *TRIM31*. The large peak on chromosome 11 seems to include two independent signals: *FSHB* (rs11031006; p = 1.08×10^−32^) and *WT1* (rs5030179; p = 5.48×10^−9^). *WT1* is located >2MB downstream of *FSHB* and the two SNPs (rs11031006 and rs5030179) are in low LD (<0.001) with each other, implying that these are two independent signals. This is further confirmed by the Locus zoom plots and FLAMES decision trees in Supplement Fig. 5 and 6 and the gene prioritization from FLAMES (Table 2). Although the *FSHB* locus was already associated with DZT in previous GWASs, *WT1* is a new finding (Table 2). *WT1* has been previously implicated in testicular cancer and ovarian cancer and is reported to play an important role in tumor invasion and metastasis.^32–34^

### Transcriptome-wide association study

To investigate functional effects of the genetic signals identified, we conducted a transcriptome-wide association study (TWAS) with FUSION.^35^ We derived expression weights from tissue that is relevant for the hypothalamic-pituitary-ovarian signaling; hypothalamus, pituitary gland, ovary, testis, vagina, breast, uterus and whole blood from GTEx v8 (Supplementary Tables 3-10; Supplementary Fig. 7-14).^36^ These analyses both confirm several previous GWAS findings and our presented new findings in the SNP-based tests. *IPO8* surpassed the tissue-specific Bonferroni correction in the hypothalamus, breast, testis and whole blood, while *SHBG* expression did so in the pituitary, and *DEAF1* expression in the hypothalamus. All three genes also were previously identified in the MAGMA or FLAMES analyses. *FSHB* was prioritized by FLAMES as the most likely gene but could not be included in the TWAS as *FSHB*’s predicted gene expression did not exhibit sufficient heritability to be considered. *ARL14EP* is located 92kb from *FSHB* and has been previously associated with other fertility traits such as PMOS/PCOS.^2,37^ A significant association was found between DZT and expression of *ARL14EP* in the pituitary gland, hypothalamus, ovary, and testis.

### Proteomic convergence of twinning and circulating *FSHB* levels

Given the *FSHB* prediction resulting from the FLAMES analysis and the well-established role of FSH in folliculogenesis, we aimed to verify the likely pathway implicated by this locus by testing the lead SNP of this locus for cis-protein quantitative trait loci (pQTL) signals. The lead SNP for DZT, rs11031006, is a fine-mapped cis-pQTL for plasma level FSHB in the FinnGen analysis of the UK Biobank Pharma Proteomics Project (p_pQTL_ = 9.5×10⁻¹⁰⁴, β_pQTL_ = -0.232, PIP = 0.54).^38^ Colocalization analysis with the GWAS locus finds a high chance of a shared causal variant (posterior probability shared causal variant (H4) = 0.997). The pQTL effect direction aligns with carriers of the minor allele having lower circulating levels of FSH and decreased incidence of DZT. The lead SNPs within the *ARL14EP* and *MMPED1* gene body also show association with circulating FSH at p = 1.01x10⁻⁷⁹ and p = 2.39x10⁻⁹ respectively. Effectively this indicates that some of the associations of DZT to *ARL14EP* and *MMPED1* in the TWAS could be the result from their association to circulating *FSHB*, rather than *ARL14EP* and *MMPED1* transcript levels.

### Heritability and polygenicity

We estimated SNP-based heritability from the updated human DZT GWAS with SBayesRC, as implemented in *gctb,*^39^ both on the complete GWAS summary statistics and on the summary statistics without genome-wide significantly associated regions (Methods) for an effective sample size of 131,893. The heritability was estimated at 3.85% (SD = 0.004), while 2.91% (SD = 0.008) heritability remained after removing the LD blocks containing the 12 significant loci from the summary statistics. This indicates that the 12 loci account for 0.94% overall heritability, which is approximately 24.4% of the total estimated SNP based heritability. The SBayesRC method also provides the so-called S estimate, which indicates the polygenicity of traits by estimating the percentage of SNPs that are expected to have a nonzero effect on a trait. The S estimate for human DZT was 0.03% (sd = 0.004%), an order of magnitude lower than for example height (2%, sd = 0.04%) or schizophrenia (5%, sd = 0.3%).^40^

### Evolutionary selection of DZT

We investigated the burden of selection of DZT in ancient DNA with the ancient genome selection (AGES) tool,^41^ on the PGS for DZT as well as the 12 leading SNPs. Overall, we show a slight negative selection on the genetic basis of DZT, as indicated by the negative genome-wide genetic correlation between the DZT and AGES summary statistics (r_g_ = -0.12, p = 0.044) (Supplementary Table 11). Of the 12 lead SNPs associated with DZT, 10 were available on the AGES website. Among these, rs11678857, located in the FSHR gene on chromosome 2, showed evidence of negative selection (Supplementary Fig. 15). Our estimates of negative selection for the DZT genes, is in line with previous estimates included in Mbarek et al (2024).^2^

### Cross-species gene-based meta-analysis implicates additional genes in twinning

To leverage independent genetic evidence from multiple species, we performed cross-species MAGMA meta-analysis, by meta-analyzing the MAGMA Z-scores of orthologous genes tagged by SNPs in both cattle and human GWAS, using the Stouffer’s Z-score method ^27,28^ (Fig. 2, Methods^42^). We compute MAGMA Z-scores for 21,361 cattle genes and 20,011 human genes. We subsequently subset to 15,184 genes mapped as orthologous with high confidence from the Ensembl database and further subsetted to 13,225 orthologous genes with a MAGMA Z-score for both humans and cattle.^43^ To verify whether we were capturing the same genetic signal we performed correlation analysis on the gene-level Z-scores. To ensure the Z-scores are on a similar scale, and have an SD of 1, Z-scores were robustly recalibrated using median-centering and MAD-based scaling. To reduce the larger noise component stemming from null-effect genes, the calibrated Z-scores were shrunk using adaptive Bayesian shrinkage to stabilize per-gene estimates. Pairwise concordance between cohorts was assessed via Pearson correlation computed on the shrunk Z-scores (Fig. 3). We found significant correlations between the two cattle GWASs (rho = 0.458; p < 1x10^-300^), and the human and cattle GWASs (human and Irish bulls: rho = 0.137, p = 8.22x10^-56^ and human and German dams: rho = 0.214; p = 2.7x10^-136^). A leave-one-chromosome-out analysis shows that this correlation is driven by genes in the FSHR peak, where leaving human chromosome 2 or cattle chromosome 11out leaves all correlations positive, but near zero and non-significant (rho = 0.007, 0.009 and 0.002), respectively. This is not entirely unexpected, given the low observed correlation (0.12) between unshrunk gene effect sizes for stature, a much more well-powered and polygenic phenotype, between cattle and humans^23^. Next, we performed attenuation weighted meta-analysis of two cattle cohorts, followed by an unweighted meta-analysis across human and cattle results (Methods).

**Figure 2.**
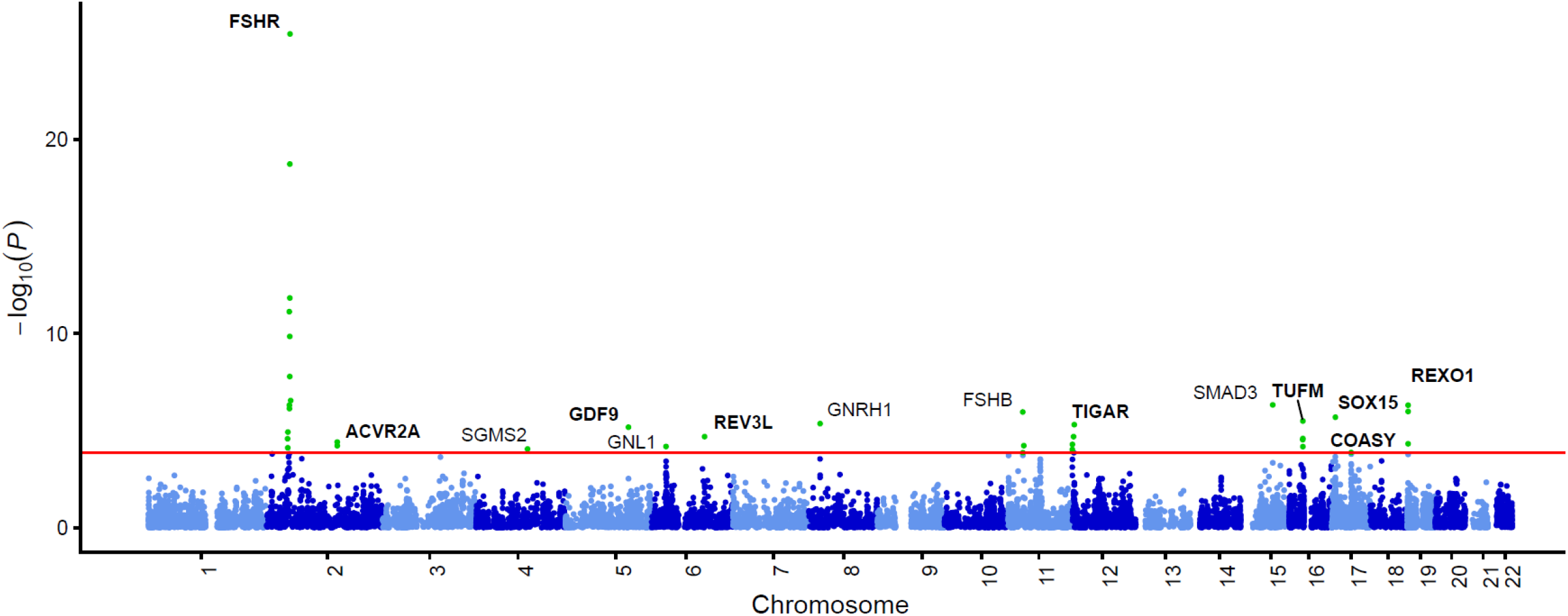
Cross-species meta-analysis gene-Manhattan. Gene Manhattan of cross-species meta-analysis of MAGMA Z-scores of human-cattle orthologous genes. Genes are projected onto the human genomic coordinates. The red line indicates FDR correction. Bold indicates that the lowest P-value is from the cross-species meta-analysis.

**Fig. 3:**
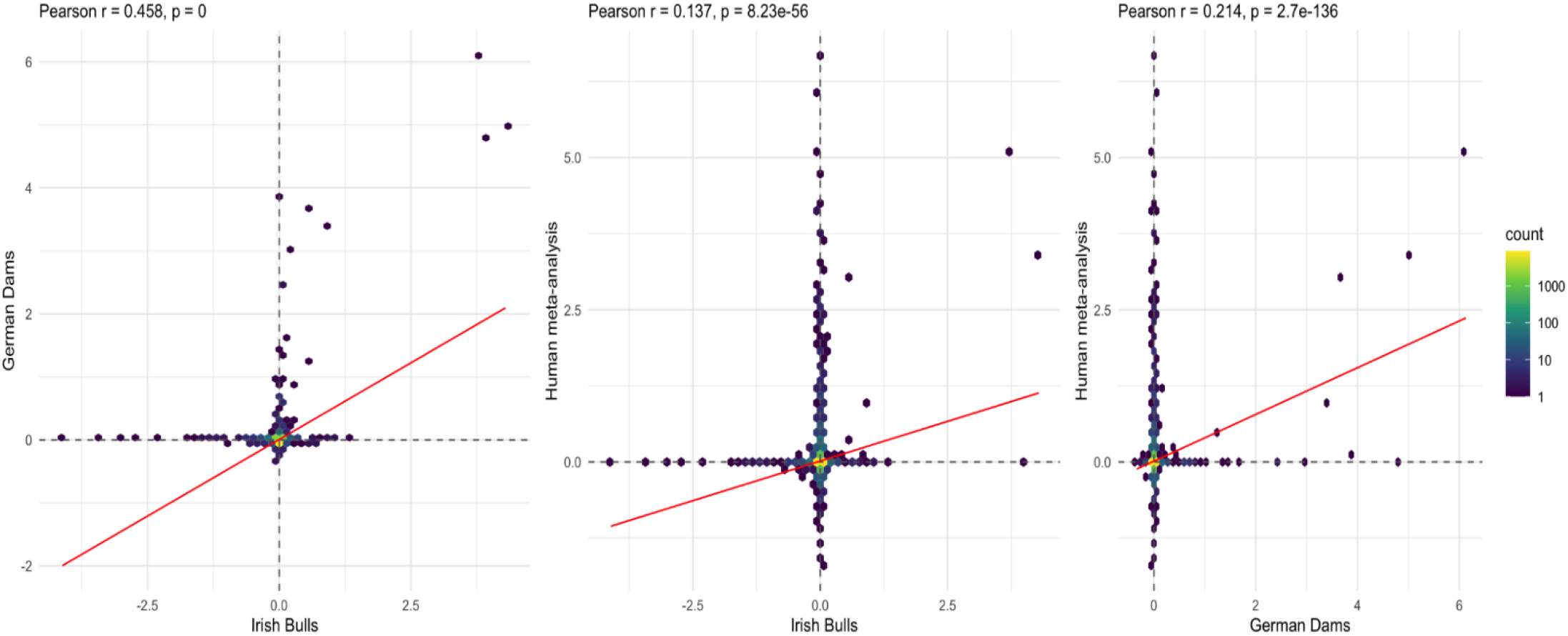
Pairwise correlation hexplots. Pairwise hexplots of cattle and human MAGMA Z-scores after adaptive Bayesian shrinkage.

In the meta-analysis, many associated loci were not common to both the human and cattle data, except for the implication of *FSHR*/*LHCGR* in DZT. Additionally, we find significant associations (after FDR correction) for 14 genes, 8 of which have not been implicated in either of the human and cattle GWAS before (Table 4). We report the genes with the highest MAGMA Z-score per locus.

**Table 4:** Genes with lowest P-value in regions below the FDR correction threshold in the gene-based meta-analyses of the human and cattle GWAS results. P-values in bold represent the lowest p-value across all analyses. Meta Z-score is equal to the sum of human and cattle Z-scores, divided by 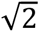.

| Gene name | Human chromosome | Cattle chromosome | Human Z-score | Human p-value | Cattle Z-score | Cattle p-value | Meta Z-score | Meta p-value |
|---|---|---|---|---|---|---|---|---|
| *FSHR* | 2 | 11 | 6.05 | 7.45E-10 | 8.82 | 5.53E-19 | **10.51** | **3.72E-26** |
| *SMAD3* | 15 | 10 | **7.03** | **1.01E-12** | -0.10 | 5.40E-01 | 4.90 | 4.75E-07 |
| *REXO1* | 19 | 7 | 4.65 | 1.67E-06 | 2.27 | 1.15E-02 | **4.89** | **4.94E-07** |
| *FSHB* | 11 | 15 | **7.66** | **9.55E-15** | -0.96 | 8.32E-01 | 4.73 | 1.11E-06 |
| *SOX15* | 17 | 19 | 3.74 | 9.26E-05 | 2.78 | 2.76E-03 | **4.61** | **2.05E-06** |
| *TUFM* | 16 | 25 | 3.62 | 1.48E-04 | 2.76 | 2.89E-03 | **4.51** | **3.23E-06** |
| *GNRH1* | 8 | 8 | **5.66** | **7.71E-09** | 0.63 | 2.64E-01 | 4.45 | 4.39E-06 |
| *TIGAR* | 12 | 5 | 2.51 | 5.96E-03 | 3.74 | 9.36E-05 | **4.41** | **4.95E-06** |
| *GDF9* | 5 | 7 | 2.22 | 1.31E-02 | 3.93 | 4.16E-05 | **4.35** | **6.68E-06** |
| *REV3L* | 6 | 9 | 3.96 | 3.68E-05 | 1.84 | 3.31E-02 | **4.10** | **2.05E-05** |
| *ACVR2A* | 2 | 2 | 2.75 | 3.02E-03 | 2.84 | 2.25E-03 | **3.96** | **3.91E-05** |
| *GNL1* | 6 | 23 | **3.96** | **3.77E-05** | 1.45 | 7.40E-02 | 3.82 | 6.62E-05 |
| *SGMS2* | 17 | 19 | **4.72** | **1.16E-06** | 0.58 | 2.81E-01 | 8.87 | 8.87E-05 |
| *COASY* | 17 | 19 | 3.21 | 6.63E-04 | 1.94 | 2.62E-02 | **3.64** | **1.35E-04** |

We find that *TUFM* reaches significance (Fig. 2; p_Cattle_ = 0.0029, p_Human_ = 0.00015, p_Meta_ = 3.2x10^-06^). This gene has previously been implicated in a consanguineous family suffering from premature ovarian insufficiency, and was subsequently validated in mouse models.^44^

*SOX15* passes the significance threshold (p_Cattle_ = 0.0028, p_Human_ = 9.3x10^-05^, p_Meta_ = 2.1x10^-06^). No previously established relationships between *SOX15* and twinning or any fertility phenotypes have been established in the literature. *SOX15* is located close to *SHBG* which is the top hit in human data and seems the more likely effector gene.

Interestingly, *GDF9* reaches significance in the meta-analysis (p_Cattle_ = 4.2x10^-05^, p_Human_ = 0.013, p_Meta_ = 6.8x10^-06^). A human genome-wide significant SNP, rs182900232, is located closely to GDF9, but GDF9 itself does not reach genome-wide significance in the human MAGMA analysis. Still, a role for *GDF9* has previously been established in DZT in humans and in sheep,^30,31,45^ and a GWAS for DZT in a Nigerian sample found one significant locus, with the lead SNP residing within the *GDF9* gene body (Hur et al., in preparation). The involvement of *GDF9* in DZT findings is consistent with the known roles of *GDF9* in promoting primordial follicle development and granulosa cell proliferation.^46^

*ACVR2A* (p_Cattle_ = 3.0x10^-03^, p_Human_ = 2.3x10^-03^, p_Meta_ = 3.9x10^-05^) plays a central role in the TGF-β pathway as one of its main receptors and in interacting with *SMAD* proteins. In mice loss of function knock out of *ACVR2A* has resulted in issues with embryonic implantation, and *ACVR2A* has been implicated for preeclampsia in humans.^46–48^

Other novel genes that surpass the significance threshold are *TIGAR* (p_Meta_ = 5.0x10^-06^)*, REV3L* (p_Meta_ = 2.1x10^-05^)*, GNL1* (p_Meta_ = 6.6x10^-05^), and *COASY* (p_Meta_ = 1.4x10^-04^) several of which have a known relation with fertility phenotypes. Loss of *TIGAR* has been implicated in meiotic defects and reduced quality of mice oocytes.^49^ *REV3L* variants have previously been associated with a GWAS for age at natural menopause.^50^ No previous association has been found between *GNL1* and twinning or reproductive phenotypes. However, given that *GNL1* is located on human chromosome 6 in the MHC region and most genes from this region are not orthologous and correlated through complex LD structure, we advise caution in interpreting this finding.

### Fine-mapping in the Cross-species analysis

Fine-mapping the likely causal variants in a locus is greatly affected by the strength of LD in that locus. Cross-species analyses can help fine-map regions likely to contain causal variants in conserved regions. Although larger haplotype regions are generally conserved, the size and fine structure of haplotype blocks differ between species.^51^ This is particularly relevant for the study of domesticated species, as their LD structures tend to be much larger as a result of selective breeding.^24^

An example of a difficult locus to fine-map is the *FSHR*/*LHCGR* locus and the cross-species analyses seem to help identify the relevant gene for DZT. The *FSHR*/*LHCGR* locus is the only region with significant GWAS variants in the separate cattle and human meta-analyses and contains genes with biologically plausible roles in DZT due to their link with the ovarian growth pathways. Fine-mapping this locus is particularly difficult in cattle, as the signal is strongest near the transcription start site of *LHCGR* but the LD block continues into the *FSHR* gene body, providing a plausible biological mechanism for both genes (Fig. 4a). The overlap between the human and cattle signal is mainly located within the *FSHR* gene body. Previous research in cattle has shown the lead SNP of the *FSHR/LHCGR* locus in cattle is an eQTL for *FSHR* expression but not for *LHCGR* expression.^17^ Combining these lines of evidence, we find that across species, the signal covers the full *FSHR* gene body; *FSHR* function strongly converges with the implication of *FSHB* as the strongest signal in the human GWAS, and eQTL follow up in cattle implicates *FSHR* even though the lead SNP in cattle is closer to the *LHCGR* transcription start side (Fig. 4). This triangulates to *FSHR* being the most likely major effector gene for DZT in the *FSHR/LHCGR* locus.

**Fig. 4:**
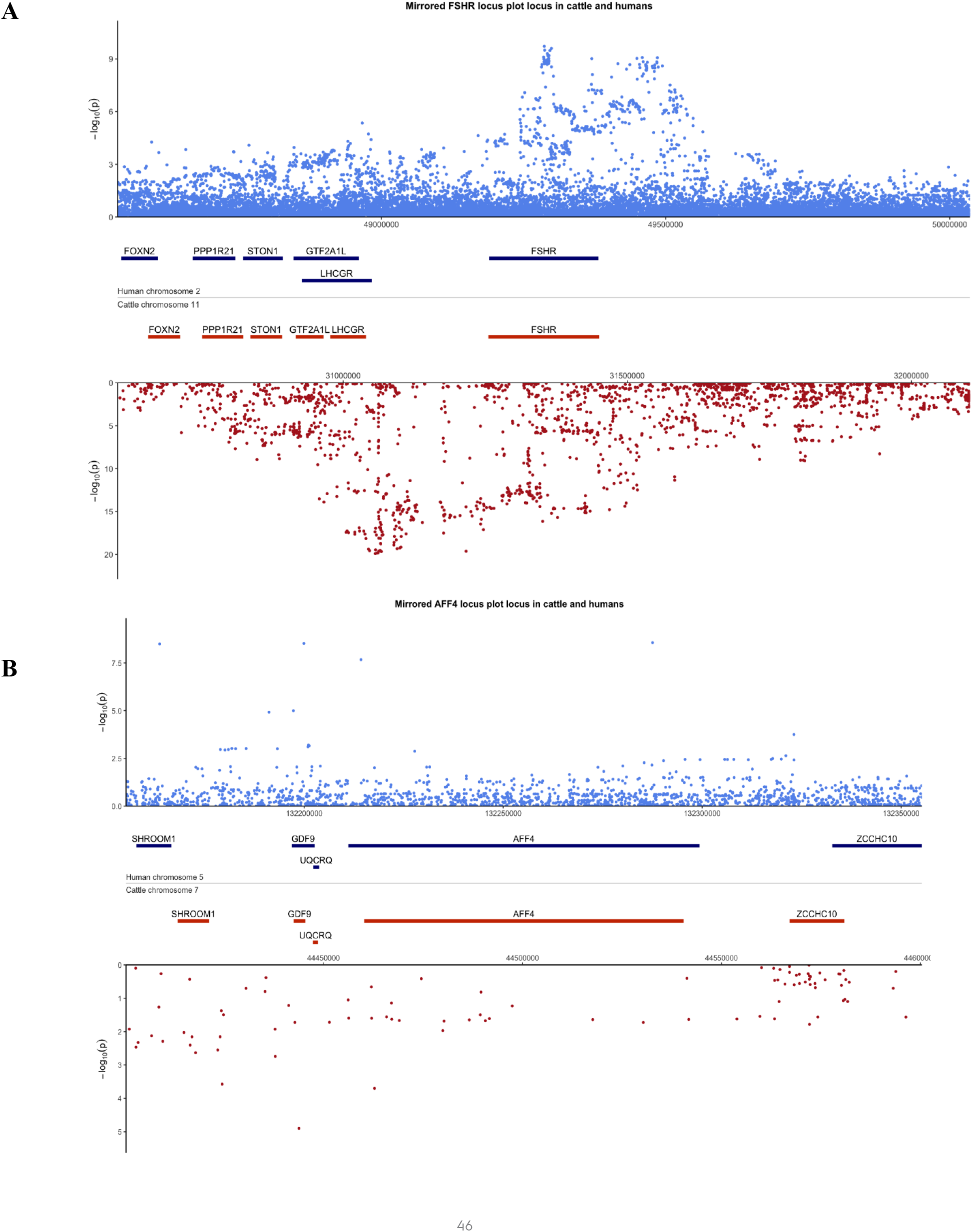
Mirrored human-cattle locus plot of the *FSHR/LHCGR* and *GDF9/AFF4* loci. Top panel (blue) shows the locus in the human genome, bottom panel (red) showing the locus in the cattle GWAS. Bottom panel contains results from the meta-analyzed cattle GWAS. Note that SNP-level cattle meta-analysis is only used for illustration in these figures. **A**) Locus centered 1.5MB around *FSHR*, **B**) locus centered 100kb around *AFF4*.

Fine-mapping is also challenging at the *GDF9/AFF4* locus. In the human GWAS, the genome-wide significant *GDF9/AFF4* locus has a lead SNP located in *AFF4* and has significant SNPs in *GDF9*. FLAMES is unable to resolve this locus, likely due to multiple strong signals being present within both the *GDF9* and *AFF4* gene bodies. Neither gene reaches significance in the human gene-based MAGMA tests while the cattle-human meta-analysis finds *GDF9* to be significant, the signal of *AFF4* nearly completely disappears. When overlaying the cattle and human locus plots, we see that in cattle the genetic signal appears to be strongest in *GDF9.* The significance in the meta-analysis and the genome-wide significant locus strongly suggest *GDF9* is the effector gene of this locus. This converges with earlier work showing rare variants in *GDF9* to be associated with DZT.^30,31^ A recently conducted GWAS in a smaller sample in Nigeria (Hur et al, in preparation), with high DZT rates, confirms this result as a SNP within the *GDF9* gene is the only genome-wide significant locus. This specific locus is undetectable in the European-ancestry samples included in this meta-analysis due to too low allele frequencies.

## Discussion

Here, we present a GWAS meta-analysis of human DZT, including data from 1,395,285 participants of European descent and a novel cross-species gene-based meta-analysis approach, investigating conserved mammalian DZT genes based on GWA meta-analyses in human and cattle. In the human GWAS meta-analysis, we identified 16 genetic regions of interest, including 12 independent loci and 4 additional genes in the gene-based analyses, and estimated a SNP-based heritability of 3.85% on the underlying scale of liability. Our results are in line with previous estimations that that DZT in humans appears to be not highly polygenic,^52^ since the 12 genome-wide significant loci explain 24.4% of the SNP based heritability and only 0.03% (sd = 0.004%) of all common SNPs are predicted to have an effect on DZT. When combining the human GWAS with cattle data in a cross-species gene-based meta-analysis framework, we identify shared genes associated with DZT. The results contain strong evidence for *FSHR* as key genes in both human and cattle and our findings of *GDF9* and *ACVR2A* further underline the influence of FSH/ovulation pathways.

The cross-species meta-analysis highlights shared, e.g. *FSHR* association in both species, as well as divergent genes, e.g. *FSHB* association in human data only, that still implicates a shared conservative biological mechanism of DZT in humans and cattle. *FSHR* is the strongest finding in the human–cattle meta-analysis. It plays a key role in folliculogenesis by encoding the receptor for follicle-stimulating hormone (FSH), a major regulator of ovarian follicle development. ^8,53^ In contrast to evidence for the FSH receptor in both species, the *FSHB* association is human specific. *FSHB* has been implicated for DZT in previous GWASs,^2,10^ as well as in several other human fertility phenotypes including PMOS/PCOS, endometriosis, and affects FSH levels in blood.^5,54–56^ The difference around *FSHB* might be due to missing effect variants in the target populations, different environments and breeding strategies / reproductive behaviors of cattle and humans, or a contrast between cattle breeds selected for production traits and an outbreeding human population. Both of *FSHR* and *FSHB,* however, point to conserved TGF-β signaling as a central pathway underlying twinning in humans and cattle. This suggests that, although the core biological process involving FSH signaling and ovulation appears to be shared across species, the relative contributions of individual genes may differ.

Our findings concerning *FSHR*, *GDF9* and *ACVR2A* further emphasize the central role of the TGF-β pathway in DZT. The TGF-β pathway has various roles in uterine processes critical for reproductive success and that influence FSH levels.^46^ *FSHR* and *ACVR2A* are both targeted receptors of this pathway while *GDF9* is one of the ligands.^46,57^ In sheep, oocyte-derived *GDF9* (growth differentiation factor 9 or FecG) is essential for normal folliculogenesis, with variants in GDF9 displaying strong associations with twinning rate. The growth factor is present in oocytes from the primary stage of follicular development until ovulation.^45,58^ *ACVR2A,* identified in the cross-species analyses, plays a central role in the TGF-β pathway as one of its main receptors and in interacting with *SMAD* proteins. Knock out mutations in mice in *ACVR2A* also show impairment of decidualization, the process of cell differentiation to support embryo implantation. These findings further support our previous hypothesis that the TGF-β pathway is central in DZT, both for influencing ovulation rate and implantation and pregnancy success.

Our cross-species gene-based GWAS meta-analysis shows two main benefits for gene discovery, gene prioritization and fine-mapping. We show that it can boost the power of gene-based analyses for genes influencing DZT liability in both species. The strongest signal for *FSHR* is observed in the meta-analysis of cattle and human data, but the locus was also identified in human and cattle analyzed separately. Importantly, the meta-analysis identifies genes at FDR significance that have moderate signal in both species, but never reach significance alone, i.e. *GDF9* from which we would infer that the gene (but not necessarily the same variants) affects DZT liability in both species.

Our method adds a new source of in gene prioritization and functional annotation data for GWAS loci: cattle eQTL data. For example, in the locus on chromosome 2 in the human GWAS, FLAMES prioritized *FSHR* as the most likely causal gene, but *LHCGR* located within the locus, also has a plausible contribution to DZT.^5^ The prioritization of *FSHR* is further supported by recent research in cattle which demonstrated that the lead SNP at the *FSHR/LHCGR* locus is an eQTL for *FSHR* expression, but not for *LHCGR* expression.^17^ In that study, granulosa cells were collected from 41 Holstein cows, and gene expression levels were tested for associations with QTL genotypes. Even though this results does not completely rule out *LHCGR,* it provides strong support for the role of *FSHR.* Comparable analysis in humans would likely rely on a less biologically relevant, but more readily available, tissue (e.g. whole blood), thereby reducing the direct interpretability of the findings.^25,36^ Our methodology therefore enables the identification of target genes shared across species, which could subsequently be investigated through QTL analyses in other species using live and biologically relevant tissues.

Our study includes limitations and challenges that should be addressed in future studies. While our analysis incorporates a large share of available human biobank data it is limited to European ancestries. Increases in diversity and sample size for human gene discovery is limited by phenotypic information on DZT and more general reproductive history in women.^59,60^ Our meta-analyses show that it is vital to remove any twin pregnancy that was conceived through assisted reproductive technology (ART) as these strongly ablate the DZT signal. In most biobanks, however, crucial information on twinning, being a twin or mode of conception often is lacking.^10,52^ It therefore remains crucial to increase the effort of collecting both genotyping and in-depth phenotyping information concerning twinning and female fertility in more and in more diverse populations. These more diverse studies are not just important for understanding DZT, but are relevant for understanding for healthy pregnancies and overall female fertility.^5,52^

We see the approach as presented in this paper as promising for comparative cross-species studies, and as a valuable addition to ongoing research in this field. Whereas we presented results based on pooling gene-level association evidence through meta-analysis, other approaches treat GWAS hits as starting points for a network-based integration. In a recent study of human obsessive-compulsive disorder (OCD) and dog compulsive disorder (OD),^61^ cross-species colocalization yielded gene communities and enriched processes that were not apparent from the, relatively small, human OCD GWAS alone, and GO enrichments were more numerous/stronger in the cross-species network than in the human-only OCD network. The method was successful, even when the implicated seed genes in the two species did not overlap.

Extending the comparative framework we have presented for DZT to other phenotypes requires several considerations. Firstly, trait definitions should be aligned at the mechanistic rather than nominal level^62^, as we demonstrate here by comparing DZT driven by multiple ovulation in human and cattle or based on careful psychometric analyses of traits as was done for human OCD and dog OD. Superficially matched labels (e.g., “depression,” “aggression”) can describe or subsume genetically distinct constructs in different species. Secondly, potential phenotypes will vary substantially in the availability of well-powered GWAS summary statistics. In cattle, genomic data exist at sufficient scale for plausible phenotypes including temperament/handling reactivity,^63–65^ whilst for other potential phenotypes such as cattle maternal separation response and human postpartum depression GWAS currently lack comparable power.^66,67^ Given these considerations, candidate phenotypes for future cross-species analysis as presented here may be prioritized according to (i) construct specificity, (ii) availability of adequately powered GWAS in both/all species, and (iii) plausibility of shared genetic architecture at the pathway level, rather than a nominal-level phenotype description.

Overall, we present the largest GWAS of DZT in humans to date, identifying 16 genetic regions of interest with a SNP-based heritability h^2^ of 3.85%. We provide a framework for cross-species gene-based GWAS meta-analysis and demonstrate its usefulness, which we leverage based on human and cattle data to strengthen evidence for human DZT loci and to identify additional genetic regions of interest. Our results support a central role for FSH/ovulation pathways influenced by the TGF-β signaling pathway for DZT in both human and cattle.

## Methods

### Participants human GWAS

We included a total of 1,395,285 participants of European descent from 10 countries across 15 samples (Table 1). Six samples performed a GWAS with mothers of twins (MODZT) as cases and seven cohorts performed a GWAS with DZ twins (DZT) as cases and two cohorts performed two GWASs in independent samples, one with MODZT as cases and one with DZT as cases (Table 1). MODZT and DZT samples excluded cases where assisted reproductive technology (ART) had been used and controls were removed if there was any knowledge of DZ twins in the family. Details on genotyping, imputation and QC are provided in Supplementary Table 13 and Supplementary data 2. Here we describe the included participants and the phenotyping.

#### NTR

The Netherlands Twin Register (NTR) provided samples for both the MODZT and DZT GWAS.^68^ The MODZT sample consisted of 1,808 cases and 6,373 controls, and the NTR DZT sample consisted of 3,810 cases and 7,695 controls. This sample also included data that was collected for the PROSPECT study cohort. The samples between the two analyses were selected to be independent of each other based on pedigree information (e.g. no overlapping families between the two samples). In case of overlap (e.g. both mother and twins genotype), priority was given to the MODZT GWAS. Zygosity was confirmed by DNA genotyping. Data on the mode of pregnancy were available from surveys sent out to mothers of twins or to parents upon registration of young twins, and from telephone interviews as part of a project on DZ twinning.^69,70^ Cases were excluded if they reported ART. If no reports on the mode of pregnancy were available, cases were excluded unless the twins were born prior to 1970.

#### QIMR

We included 4,942 cases and 29,327 controls from Australia and New Zealand. These data were collected in two waves: the previously analyzed sample of 3,273 cases and 24,009 controls (ANZ2023)^71^, and the newly collected sample of 1,669 MODZT cases and 5,318 controls (ANZ2026)^72^. Cases were screened for the use of fertility treatments based on questionnaire data.

#### MCTFR

All subjects in this sample were identified through Minnesota state birth records and recruited to establish a population-based registry of twins at the Minnesota Center for Twin and Family Research (MCFTR).^73^ The current study included 568 mothers of DZ twins and 1,862 control subjects who were the parents of MZ twins from 1,062 families, including 800 complete parental pairs, 203 mothers, and 59 fathers. Most twins were born in the 1970s or early 1980s, when only few had access to fertility treatments.

#### WGHS

The Women’s Genome Health Study (WGHS) is a population-based, prospective cohort of initially healthy, female North American health care professionals at least 45 years old at baseline, representing participants in the Women’s Health Study (WHS) who provided a blood sample at baseline and consented for blood-based analyses.^74^ The WGHS cohort has been described previously.^74^ Additional information related to health and lifestyle were collected by questionnaire throughout the WHS trial and continuing observational follow-up. A total of 361 mothers of DZ twins and 20,183 controls from the WGHS were included in the current study. MODZT were of reproductive age before the common use of ART (born at the latest in 1951).

#### deCODE

Mothers of twins or other multiples (‘twins’) were selected from among those taking part in deCODE’s genetic studies based on a nation-wide genealogical database.^75^ Control subjects were individuals participating in deCODE’s genetic research, from which both mothers of twins and the mothers’ first-degree relatives had been removed. In total, the deCODE sample included in the current study consisted of 2,877 mothers of DZ twins and 212, 767 controls. Only mothers who had given birth before 1991 were included, to ensure no fertility treatments were used.

#### FinnGen

In total, 2,009 MODZT were included as cases and 478,349 participants as controls for MODZT GWAS from the FinnGen database release 12 (R12).^76^ Cases include mothers with sex-discordant twins, excluding births involving assisted reproductive technologies. Mothers of twins were identified based on birth and population registry data. Analysis includes only sex-discordant twins to ensure zygosity, as complete information on zygosity was not available. Mothers of twins with triplets or more children from a single delivery were excluded from the study. 1st and 2nd degree relatives of MODZT, and participants aged <17 yrs. were excluded from controls. Samples from the Finnish twin study were excluded.

#### MoBa

The Norwegian Mother, Father and Child Cohort Study (MoBa) is a population-based pregnancy cohort study conducted by the Norwegian Institute of Public Health.^77^ MoBa contributed 658 MODZT and 53,046 controls and 474 DZT and 40,622 controls to the meta-analyses efforts. The two analyses contained independent cases and controls. Data on the participating mothers were linked with data from the Medical Birth Registry of Norway (MBRN; a national health registry containing information about all births in Norway) to assess their reproductive history. Mode of conception was reported by record linkage to the MBRN, which contains information on IVF/ICSI use as reported from the fertility clinics, or by self-report in MoBa.^70,78^ Mothers who received fertility treatments were excluded from the sample.

#### TwinLife

We included 331 MODZT cases and 660 controls from the TwinLife cohort.^79^ The information about the mode of twins conception was based on maternal self-report on mode of conception (no medical support). The mean year of birth of mothers of the twins included in the GWAS was 1970, being all of them between 1951 and 1989. The genotype data used in the present study was quality-controlled and imputed together with the full genotyped subset of the TwinLife cohort. For a more detailed description of the TwinLife genotype data processing, see Deppe et al. (2024) and Pahnke et al. (in-prep).^80^ Briefly, DNA was extracted from saliva samples and genotype data was generated using Global Screening Arrays (GSA+MD-24v3.0-Psych-24v1.1, Illumina, San Diego, CA, USA). SNPs with p < 1e-6 for HWE, MAF < 0.001 and a call rate <98% were removed pre-imputation. Samples were removed if they were duplicated, were flagged as a sex and/or family-mismatch, had undetermined sex, a call rate <98%, autosomal heterozygosity deviation (FHET) within +/-0.20, were PC outliers, defined as samples whose PC values deviated more than 6 standard deviations from the mean of the first 10 PCs, were not assigned to European superpopulation as defined by 1000 Genomes reference The imputation was performed using 1000 Genomes phase 3^81^ as the reference panel. Summary Statistics were unfiltered for r^2 and non-snp variants like indels.

For all TwinLife participants informed consent was collected (including parental consent for minors). TwinLife was ethically approved by the Ethics Committee of the German Psychological Society (DGPs; protocol number: RR 11.2009). The molecular genetic analyses were approved by the Ethics Committee of the Medical Faculty of the University of Bonn (No. 113/18).

#### STR

A total of 15,251 DZ (cases) and 3,420 MZ twins (as controls, one per pair) were included in the analysis from the Swedish Twin Registry (STR).^82^ Three sub-studies of the Swedish Twin Registry, denoted TwinGene, SALTY, and STAGE, were used. All included twins were restricted to the year of birth < 1970 to ensure naturally conceived pregnancies.

#### UK Biobank

We analyzed data from the United Kingdom Biobank (UKB) release 2. The UKB cohort contains data for 488,377 participants from across the UK, aged between 40 and 69 years, collected between 2006 and 2010. The data on twinning came from the item: ‘Are you a twin, triplet or other multiple birth?’ (UKB questionnaire field ID 1777). The options were: ‘Yes’, ‘No’, ‘Do not know’, and ‘Prefer not to answer’. We focused on 8,962 participants with Caucasian ancestry who reported being part of a multiple birth and who could not be identified as a MZ twin, and 409,591 controls. Detailed information on the selection of twins can be found in Mbarek et al. (2019).^83^

#### FTC

The Finnish Twin Cohort (FTC) study comprises three longitudinal studies of twins born before 1958,^84^ twins born 1975–1979,^85^ and twins born 1983–1987.^86^ A total of 8,188 twins were included in the analyses (6,021 DZ twin individuals and 2,167 MZ twin individuals used as controls).

#### TEDS

The Twins Early Development Study (TEDS) is a longitudinal twin study that recruited over 16,000 twin pairs born between 1994 and 1996 in England and Wales through national birth records.^87^ The sample consisted of 2,919 DZ twins and 2,580 MZ twins (one per pair), as controls were included from TEDS UK. The information gathering on the mode of the twins’ conception was based on a questionnaire at first contact with mothers regarding whether they became pregnant using fertility treatment or not.

#### Twins UK

The TwinsUK sample of MZ and DZ twin pairs was drawn from the TwinsUK adult twin registry, a cohort designed to study the heritability and genetics of age-related diseases. The cohort has been recruited from the general population through national media campaigns in the UK and is comparable to age-matched population singletons in terms of clinical phenotype and lifestyle characteristics.^88^ A total of 5,697 twins (2,206 DZ twin individuals and 3,491 MZ twin individuals as controls) from TwinsUK were included. Twins were restricted to the group for which we had verified information confirming they were conceived without the use of fertility treatments.

#### DTR

A total of 1,186 DZ twins and 391 MZ twin controls were included in the analyses from the Danish twin register (DTR).^89^ To ensure natural conception, only twins born before 1970 were included.

#### Lifelines

Lifelines is a multi-disciplinary prospective population-based cohort study examining in a unique three-generation design the health and health-related behaviours of 167,729 persons living in the North of the Netherlands. It employs a broad range of investigative procedures in assessing the biomedical, socio-demographic, behavioral, physical and psychological factors which contribute to the health and disease of the general population, with a special focus on multi-morbidity and complex genetics.^90^ Lifelines contributed 718 DZ twins and 67,657 controls. Controls consist of MZ twins, their families and singletons’ families. Information available on the mode of twins’ conception was based on maternal self-reporting or twin reports on the mode of conception, and on the year of birth for twins before 1970.

### GWAS and Meta-analyses

A genome-wide association analysis was conducted in each participating cohort by logistic regression under an additive genetic model using fastGWA-BB.^91^ Analyses were adjusted for population structure using the first ten principal components (PC) of genetic ancestry and study-specific factors such as batch and genotyping platform and the genetic relatedness matrices (GRM) as covariates. For complete MZ twin pairs, one twin per pair was randomly selected. Before meta-analysis, we removed SNPs with low minor allele frequencies (<0.0001), low imputation quality (r^2^ < 0.8) and deviations from Hardy–Weinberg equilibrium (P < 10^−5^) from each cohort. A total of 28,162,584 variants passed these quality control steps and were kept in the meta-analysis. Next, we collated the individual GWAS results through meta-analysis. Results from individual GWAS (MODZT and DZT) were combined by the fixed-effects inverse variance method based on the regression β estimates and standard errors from each study implemented in METAL software.^92^ DZ twins carry half of the genetic information of MODZT. Genotyped DZ twins for whom no maternal genotype data were available were included in the meta-analysis, taking account of the differential genetic information from mothers of twins and twin offspring as described in the supplementary data of Mbarek et al 2024.^2^ Briefly, the regression coefficient from the DZ twin GWAS is multiplied by 2 to convert it to the same scale as the coefficient from the MODZT GWAS and the standard error is scaled accordingly using the delta method, whereby multiplying the coefficient by 2 also multiplies its standard error by 2, allowing the two estimates to be combined in the meta-analysis. The genome-wide significance threshold was set at the level of P < 5.0×10^−8^.

Functional Mapping and Annotation (FUMA) was used to provide genome-wide, gene-wide and regional visualization of the results.^93^ Before the results were visualized by FUMA, we filtered SNPs on effect allele frequency (EAF) <0.01 and SNPs had to be measured in at least the mean (561,706) of participants included per SNP, retaining 11,370,680 SNPs. Full details of participants, cohorts and genotyping can be found in Table 1 and Supplementary Table 8. Full details of the individual GWASs and the meta-analysis plan can be found in the standard operating procedure circulated to the consortium study cohorts and in Supplementary Data 1, as well as the earlier papers of the TGC.^2,10,83^

### Fine-mapping and Gene prioritization

For fine-mapping, FUMA was employed to identify the independent significant SNPs that showed a genetic signal.^93^ Multi-marker Analysis of GenoMic Annotation (MAGMA) analysis was performed to run the gene-based test and to look at tissue enrichments.^26^ In MAGMA, input SNPs were mapped to 19,171 protein coding genes, so genome-wide significance was defined at P = 0.05/19,171 = 2.61 × 10^−6^. MAGMA was run with the standard settings. Gene prioritization software, fine-mapped locus assessment model of effector genes (FLAMES),^29^ was used to identify the most likely causal genes for each of the independent SNPs.

FLAMES utilizes the standard output of FUMA and MAGMA as starting points for the analyses and creates machine learning predictions from biological data linking SNPs to genes. It then evaluates these scores together with gene-centric evidence of convergence of the GWAS signal in functional networks. All six steps of FLAMES were performed as described in the documentation (https://github.com/Marijn-Schipper/FLAMES/tree/master). The FLAMES score of a gene denotes the combined evidence of functional convergence with other GWAS-implicated genes and the evidence from biological studies that link genes to fine-mapped SNPs in the locus of interest. The inputs for the classifier are created with the FLAMES annotation step, which annotates a credible set with various SNP-to-gene annotations. Credible sets were generated by fine-mapping the FUMA loci with the PolyFun implementation of SuSiE,^94,95^ under a single causal variant assumption. This single-variant assumption might not be true for each locus. Under the single-causal variant assumption, in the presence of multiple causal variants, depending on their strength of association, the signals will be aggregated into one credible set on which to make a FLAMES call on. However, given the complex LD association pattern of the meta-analysis and data-sharing preventing from using in-sample LD, multiple-causal variant fine-mapping is unlikely to be reasonably calibrated.^96^ Network convergence-based PoPS scores were generated using the features provided by the FLAMES study, using the previously described MAGMA scores as input. The convergence-based scores and the locus-based SNP-to-gene scores are scaled and combined into a final FLAMES score for each gene in the locus. We prioritized genes above the recommended FLAMES threshold of a scaled FLAMES score larger than 0.248 and a raw FLAMES score larger than 0.136, ensuring that the set of predicted genes will cumulatively have an estimated precision of at least 75%.

### Heritability

SNP-based heritability was estimated using SBayesRC, which is part of the *gctb* software.^39,97^ The results were first filtered on the effect allele frequency (EAF) of 0.01 ≤ EAF ≤ 0.99. Variant EAF and the effect sizes were aligned with the NTR reference for the 1000 genomes variants. Discovery variants that were not part of this reference were discarded. Two heritability estimates were obtained: 1) based on all SNPs included in the meta-analyses, and 2) based on all genome-wide SNPs minus the independent significant SNPs and SNPs in LD with the significant SNPs. All SNPs within a 200kb window were removed. The independent significant SNPs were based on the information provided by FUMA. The heritability was estimated on the liability scale, with an estimated prevalence of 1%. For the effective sample size was calculated using (4 x cases x controls)/(cases + controls), whereby the DZT counted as half cases, leading to an effective sample size of 131,893.

### Transcriptome-wide association study

We conducted FUSION TWAS^35^ applied to the human meta-analyzed GWAS summary statistics to identify genes whose cis-regulated, predicted gene expression is associated with DZT. We used FUSION expression weights derived from hypothalamus, pituitary gland, ovary, testis, vagina, breast, uterus and whole blood GTExv8 tissue libraries^25,98^ of European genetic ancestry. We used the 1000 Genomes European linkage disequilibrium reference panel for all analyses. FUSION weights were only included for genes whose expression exhibited significant heritability by Genome-wide Complex Trait Analysis (GCTA). Within-tissue Bonferroni-correction was used to establish TWAS p-value significance and was based on the number of TWAS p-values generated per tissue. Genes within the major histocompatibility complex were not included, due to extensive linkage disequilibrium.

### FSHB colocalization analysis

We performed approximate Bayes Factor (ABF) colocalization analysis between our human DZT meta-analysis summary statistic and the Sun *et al*. analysis of plasma FSHB protein levels. Specifically, we matched overlapping variants on their position on GRCh37 if the fall in the region from chr11

### AGES

Utilizing the Ancient GEnome Selection (AGES) summary statistics by Akbari et al (2026),^41^ we assessed overall selection on the genetic basis of DZT over the past 10,000 years. Specifically, we used linkage disequilibrium score regression (LDSC)^99^ to estimate genome-wide genetic correlations between DZT effect sizes and AGES-derived selection coefficients. We additionally performed a look up of the leading SNPs on the AGES website (https://reich-ages.rc.hms.harvard.edu/#/).

### Cattle German Holstein GWAS

We performed a GWAS of 86,391 German Holstein dams, of which 3,094 had given birth to twins. Information on the zygosity of the twin offspring was not available, however, the majority are expected to be DZ twins as only 5% of all twin births in cattle are monozygotic.^100^ Micro-array data were imputed as described in Križanac, et al. (2025), leaving to 3,393,224 SNPs after filtering to a minimum minor allele frequency of 0.01.^101^ Given the low prevalence of the binary phenotype (3.58% in this sample) and high relatedness among samples, the GWAS analysis was performed using SAIGE.^102^ SAIGE uses saddle point approximation to deal with low-prevalence binary phenotypes and constructs a GRM to deal with relatedness. The null model was constructed using 57,926 LD-independent SNPs, adding a binary covariate to account for the use of sexed semen during insemination, a categorical covariate to account for the year and season of the recorded birth, and the first 10 genetic PCs calculated on the whole set of SNPs, to account for population structure. The resulting GWAS seems to be well calibrated, with genomic inflation estimated to be *λ_GC_*= 0.99 (Supplementary Fig. 16).

### Cattle Irish Holstein GWAS

We included the GWAS summary statistics from Kirkpatrick et al., where GWAS was performed on the estimates of genetic merit for twinning from 2,656 Holstein-Friesian sires.^7^ Genetic merit per bull was calculated based on the twinning occurrences in the daughters of these bulls, from a total of 6,748,725 calving records from 2,238,936 cows.

### Cross-species gene-based meta-analysis

We performed MAGMA gene-based analysis on the human and two cattle GWAS. Human MAGMA analysis was performed using FUMA, using the European 1000Genomes individuals as a reference panel. For all three MAGMA analyses we used a window of 0, meaning only SNPs within the gene body were annotated to genes, and used the default --snp-wise=mean model. Cattle gene annotations were generated using the Ensembl (v.110) gene annotations of the bovine ARS-UCD1.2 reference genome. MAGMA^26^ gene-based testing was performed using Holstein cattle included in the run8 1000Bulls reference genotypes^103^ for the Irish Holsteins,^104^ for which only summary statistics were available. The summary statistics of the 86,391 German Holsteins were analyzed using the full in-sample PLINK files as the LD reference.^105^

We meta-analyzed the cattle MAGMA Z-scores using Stouffer’s Z-score method^106^. Although we also performed SNP-level inverse-variance weighted meta-analysis between the two cohorts for plotting, it is preferable to run MAGMA on the two cohorts separately, and meta-analyze the gene-based results due to the better matching of LD reference panels. Weighted Stouffer’s Z should improve statistical efficiency and power relative to an unweighted meta-analysis,^28^ but proper inverse-variance weighting (IVW) is not achievable here. First, postGSf90 imposes MAF-dependent shrinkage on effect sizes, so the reported standard errors do not have the same scale interpretation as frequentist standard errors from SAIGE.^107^ Second, postGSf90’s standard errors themselves rely on a diagonal approximation of the prediction error variance, which is a further departure from the properties assumed by IVW. Together, these mean the postGSf90 and SAIGE SEs are not comparable variance estimates and cannot be combined directly. A sample-size-based weighting scheme is similarly not viable: the cattle and human phenotypes differ in scale (quantitative vs. case-control), and relatedness among the cattle samples further complicates any effective sample size calculation. Instead, to obtain an approximate measure of relative precision between the two GWAS, given the reasonable assumption that both studies are measuring the same underlying phenotype and identify SNPs that reach genome-wide significance in both, we calculate the attenuation coefficient between the Z-scores of variants that are significant in both studies (p < 5e-8). We use the median as an empirical attenuation factor to weight the studies in the gene-based meta-analysis. The attenuation factor of 1.1 gives that the German Holstein Dams are upweighted compared to the Irish Holstein Bulls. As a sensitivity analysis, relaxing the threshold to nominally significant SNPs (p < 0.05) yielded a similar coefficient (1.102 instead of 1.1). The cattle Z-scores for both respective studies are calculated as:

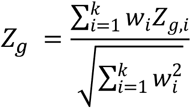

Where:

*Z_g_* is the overall combined meta-analytic Z-score for gene *g*.

*Z_g_*_,*i*_ is the MAGMA Z-score for gene *g* for the *i*-th cattle cohort.

*w_i_* is the weight assigned to cattle cohort *i* based on SNP-level attenuation analysis.

Meta-analysis of the cattle and human MAGMA Z-scores was performed using the unweighted Stouffer’s Z method.^27,28^ Given that in a cross-species meta-analysis the species are the unit of replication, sample-size weighting is not appropriate.^28^ Practically, the meta analyzed Z-scores are therefore:

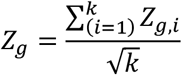

Where:

*Z_g_* is the overall combined meta-analytic Z-score for gene *g*.

*Z_g_*_,*i*_ is the MAGMA Z-score for gene *g* for the *i*-th species.

*k* is the total number of independent studies being combined.

Human-cattle meta-analysis was restricted to cattle-human orthologous genes. We mapped cattle-human orthologs using the Ensembl predicted orthologous genes, keeping only genes that have the maximum orthology score (1 in the range of 0-1), resulting in 13,225 orthologous genes tagged by the combined GWAS. Given the exploratory nature of this analysis we report significant genes at a 5% Benjamini-Hochberg FDR threshold.

## Acknowledgements

We warmly thank all participants of the NTR and all previous employees who contributed to the data collection and preparation. We thank the Australian Multiple Birth Association (AMBA) in Australia, and NZMBA, Allison Miller, Joy Brown and Carolyn Lister in NZ for support with sample collection and related processes. We thank Prof David Whiteman and A.Prof Catherine Olsen for access to the genotyping and survey data for participants in their QSkin and QSkin-II studies, who were used as controls for ANZ2023 and ANZ2026 respectively. We thank all FinnGen participants, principal investigators, laboratory personnel and data management teams. We warmly thank all participants of MoBa and all previous employees who contributed to the data collection and preparation. Swedish Twin Registry is acknowledged for access to data. The Swedish Twin Registry is managed by Karolinska Institutet and receives funding through the Swedish Research Council under the grant no 2021-00180. We gratefully acknowledge the ongoing contribution of the participants in the Twins Early Development Study (TEDS) and their families. We acknowledge the access to the bonna cluster at the University of Bonn and the support provided by the HPC@HRZ team. We thank Shirin Zare for performing the DNA extraction. The authors wish to acknowledge the services of the Lifelines Cohort Study, the contributing research centers delivering data to Lifelines, and all the study participants.

## Data availability

The results from the human GWAS MA will be made available through the GWAS catalog. The SOP of the Twinning Genetic consortium is available as supplementary information and at the Github repository (https://github.com/hubersn1998/DZT_GWAMA3). Other scripts that were used for the main analyses are also available on the github repository.

The Finnish biobank data can be accessed through the Fingenious® services (https://site.fingenious.fi/en/) managed by FINBB. T.T. was funded by Sigrid Jusélius Foundation (https://sigridjuselius.fi/en/), the HiLIFE Fellows Program, and the Research Council of Finland (grant number 315589).

Data from the MoBa and the MBRN used in this study are managed by the national health register holders in Norway (Norwegian Institute of public health) and can be made available to researchers, provided approval from the Regional Committees for Medical and Health Research Ethics (REC), compliance with the EU General Data Protection Regulation (GDPR) and approval from the data owners. The consent given by the participants does allow storage of data on an individual level in repositories or journals. Researchers who want access to data sets for replication should apply through www.helsedata.no. Access to data sets requires approval from The Regional Committee for Medical and Health Research Ethics in Norway and an agreement with MoBa.

## Author contributions

NH, MS, JJH, HM, GW, GWM, BWK, CBL. NGM and DIB contributed to the conceptualization of the study. NH, MS, SGo, MB, IMN, PvdM, RP, JvD, Jb, EE, EV, KC, MR, CM, CK, AJF, MK, HS, JK, CS, LH, DPB and BWK prepared the data for the analysis. NH, MS, SGo, JL, ÁG, CMP, MN, CKLP, RK, TP, JMB, TRM, KK, WL, EH, MKN, SGa, AG, SLPS, CS, LH, DBP and BWK contributed to the analyses of the study. NH, MS, JMB, TRM, VFM, contributed to the visualization of the study. NH, JJH, HM contributed to setting up the GWAS meta-analysis pipeline. NH, MS and DIB drafted the first version of the manuscript. YM, JvD, CdL, YMH, BR, VM, CDS, HS, KC, FW, RP, MO, PKEM, CK, AJF, CCM, DIC, WI, CBL, JRH, TT, MK, HS, JK, GWM, BWK, GW, NGM and DIB provided supervision. All authors provided feedback on the manuscript and support the submitted version of the manuscript.

