## Supplementary Figures for "Combined genome-wide association studies in humans and cattle: Cross-species genetic insights into dizygotic twinning"

**Supplementary Fig. 1:** QQplot of the A) SNP-based GWAS meta-analyses using METAL and B) the gene-based MAGMA analyses.

**Supplementary Fig. 2:** Locus plot and FLAMES decision plot for chromosome 5 rs182900232

**Supplementary Fig. 3:** Locus plot and FLAMES decision plot chromosome 6 rs1611747 and rs1362104

**Supplementary Fig. 4:** Locus plot and FLAMES decision plot chromosome 11 rs4074376

**Supplementary Fig. 5:** Locus plot and FLAMES decision plot chromosome 11 rs11031006

**Supplementary Fig. 6:** Locus plot and FLAMES decision plot chromosome 11 rs5030179

**Supplementary Fig. 7:** Results of the transcriptome-wide association study (TWAS) using FUSION[29] in the Pituitary

**Supplementary Fig. 8:** Results of the transcriptome-wide association study (TWAS) using FUSION[29] in the Hypothalamus

**Supplementary Fig. 9:** Results of the transcriptome-wide association study (TWAS) using FUSION[29] in Whole Blood

**Supplementary Fig. 10:** Results of the transcriptome-wide association study (TWAS) using FUSION[29] in the Ovary

**Supplementary Fig. 11:** Results of the transcriptome-wide association study (TWAS) using FUSION[29] in the Testis

**Supplementary Fig. 12:** Results of the transcriptome-wide association study (TWAS) using FUSION[29] in the Vagina

**Supplementary Fig. 13:** Results of the transcriptome-wide association study (TWAS) using FUSION[29] in Breast tissue

**Supplementary Fig. 14:** Results of the transcriptome-wide association study (TWAS) using FUSION[29] in the Uterus

**Supplementary Fig. 15:** The burden of selection of DZT in ancient DNA with the ancient genome selection (AGES)[35] for rs11678857

**Supplementary Fig. 16:** Cattle GWAS Q-Q plot.

**Supplementary Fig. 1:** QQplot of the A) SNP-based GWAS meta-analyses using METAL and B) the gene-based MAGMA analyses.

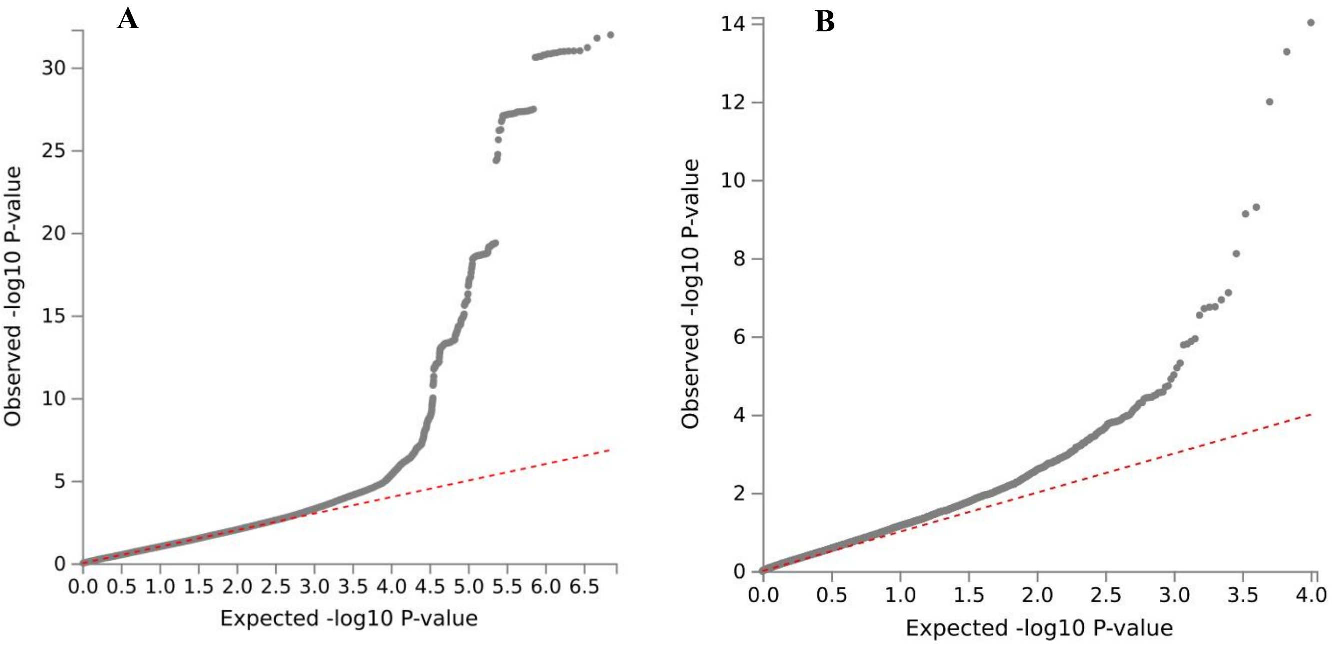

**Supplementary Fig. 2:** Locus plot and FLAMES decision plot for chromosome 5 rs182900232

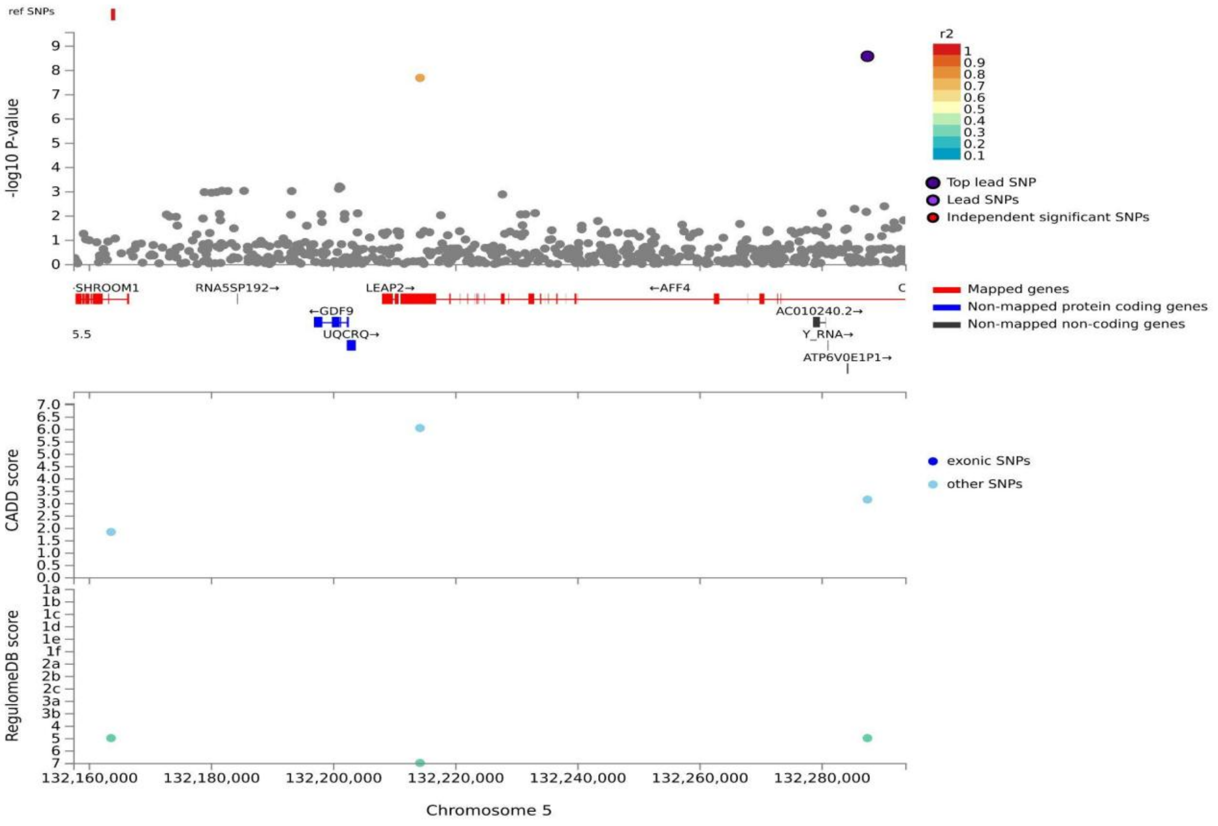


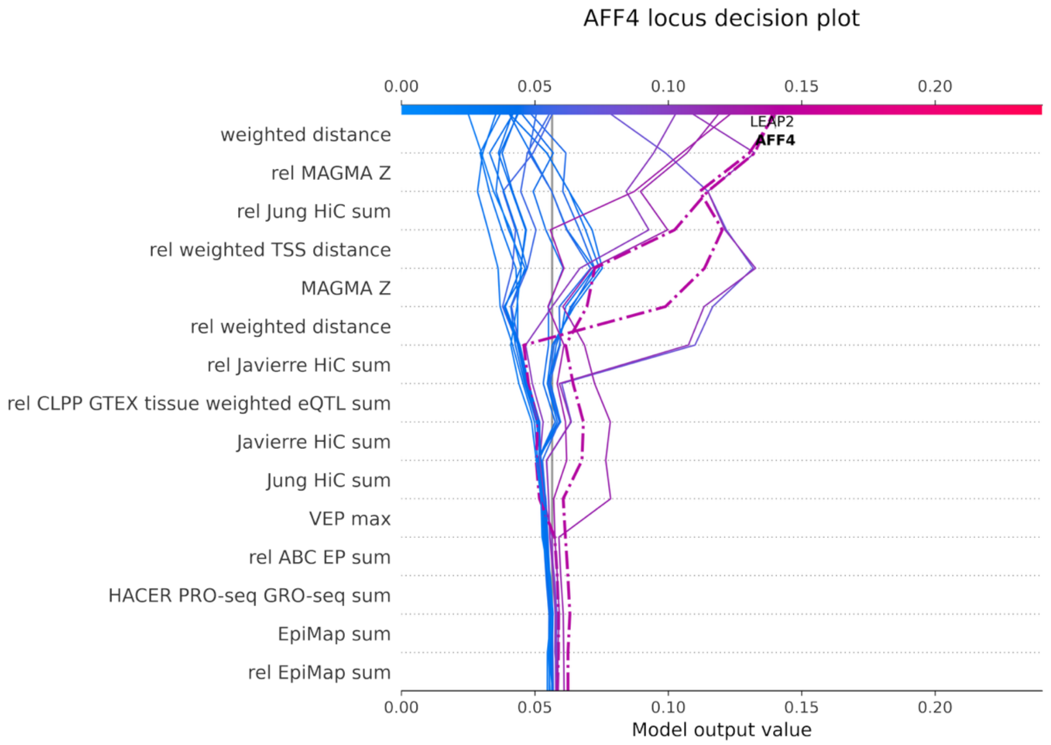

**Supplementary Fig. 3:** Locus plot and FLAMES decision plot chromosome 6 rs1611747 and rs1362104

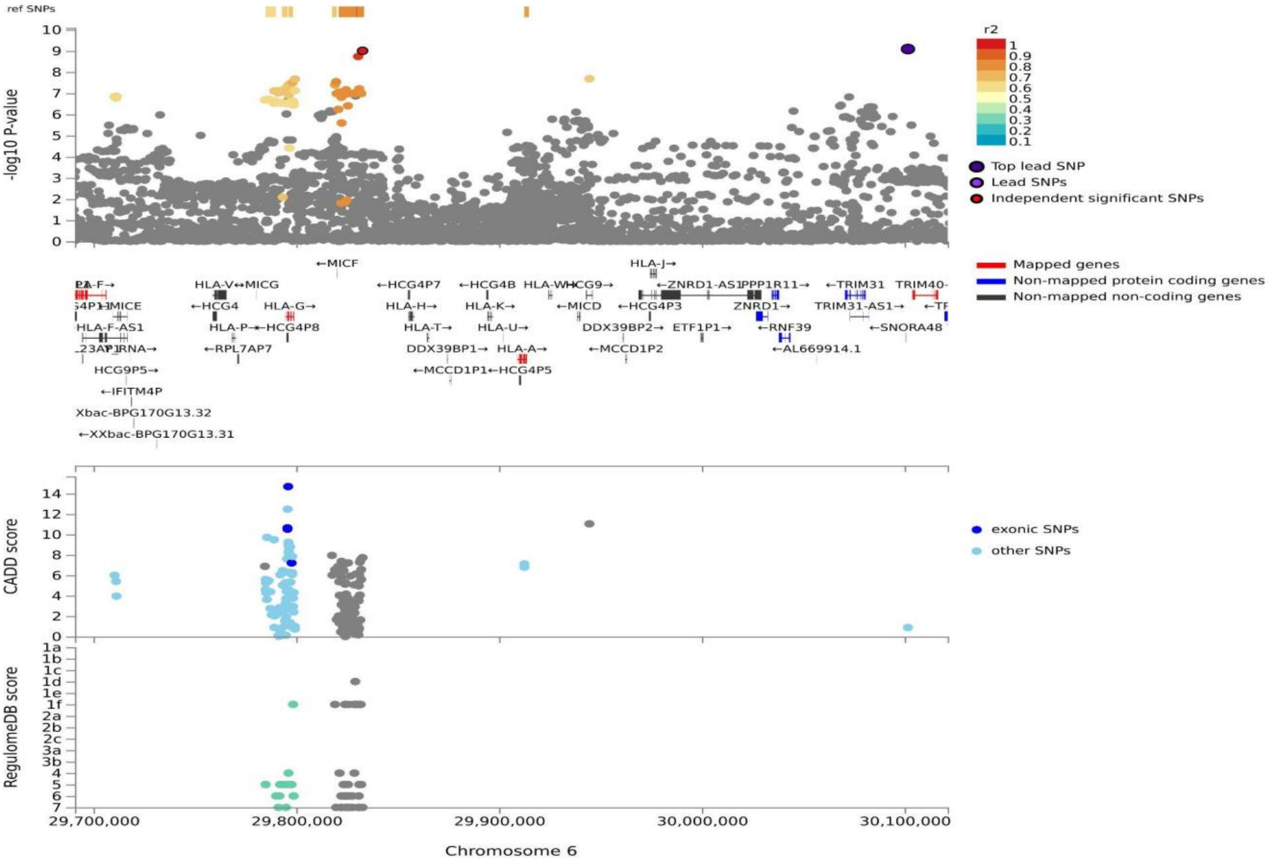


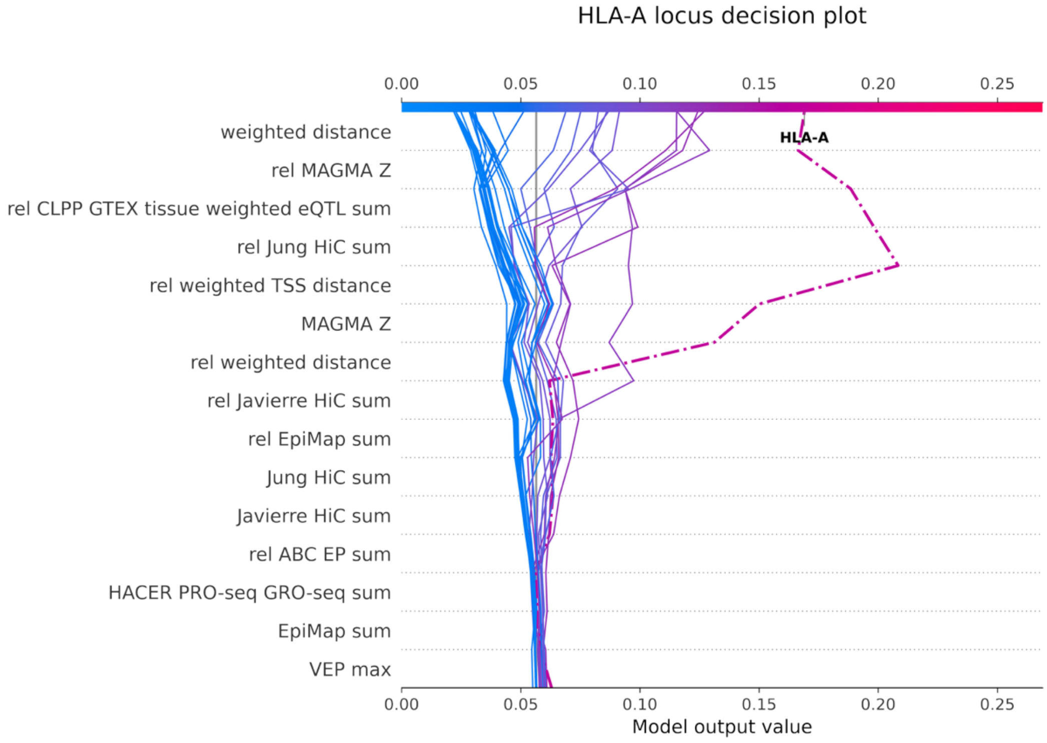

**Supplementary Fig. 4:** Locus plot and FLAMES decision plot chromosome 11 rs4074376

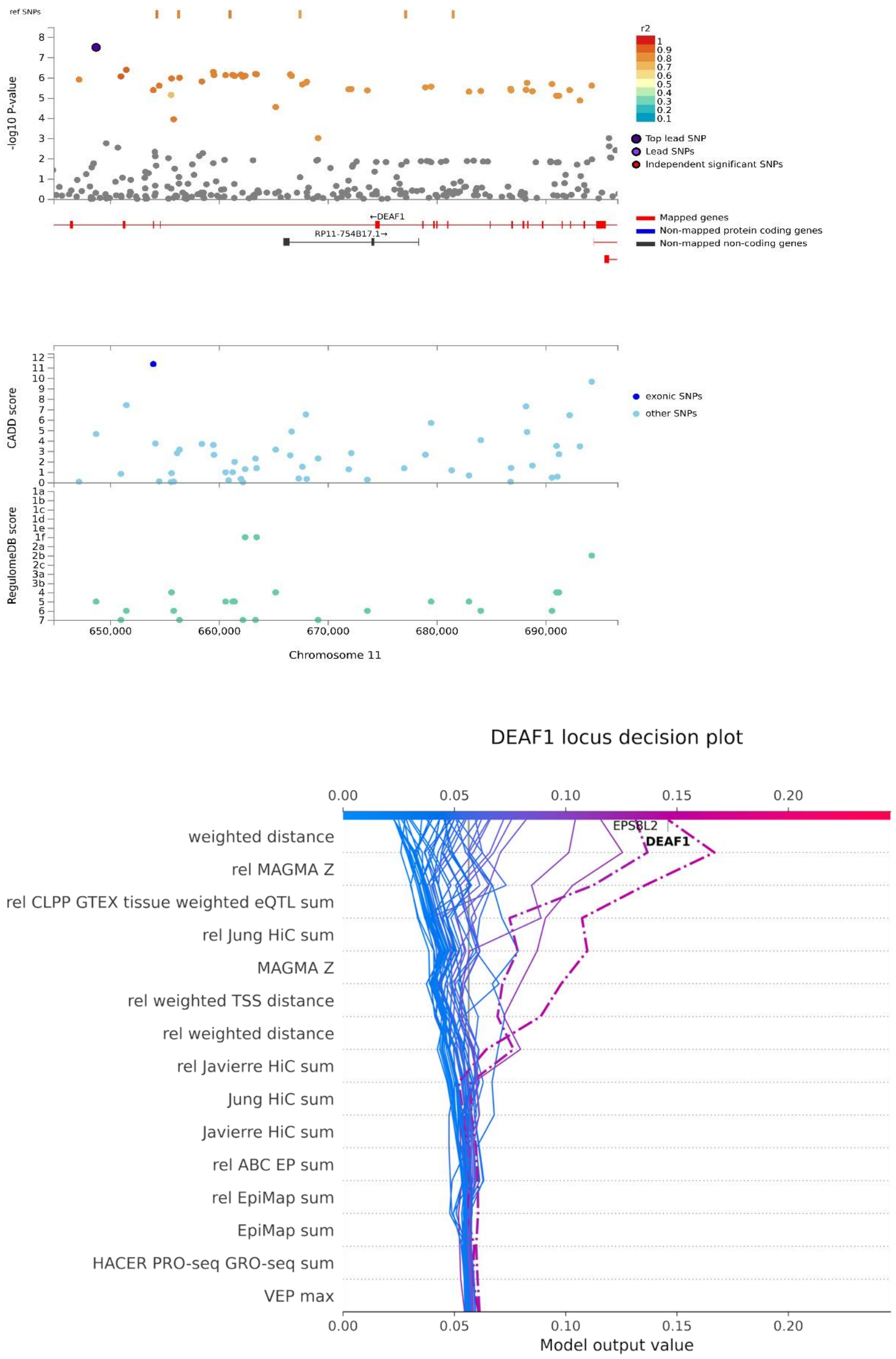

**Supplementary Fig. 5:** Locus plot and FLAMES decision plot chromosome 11 rs11031006

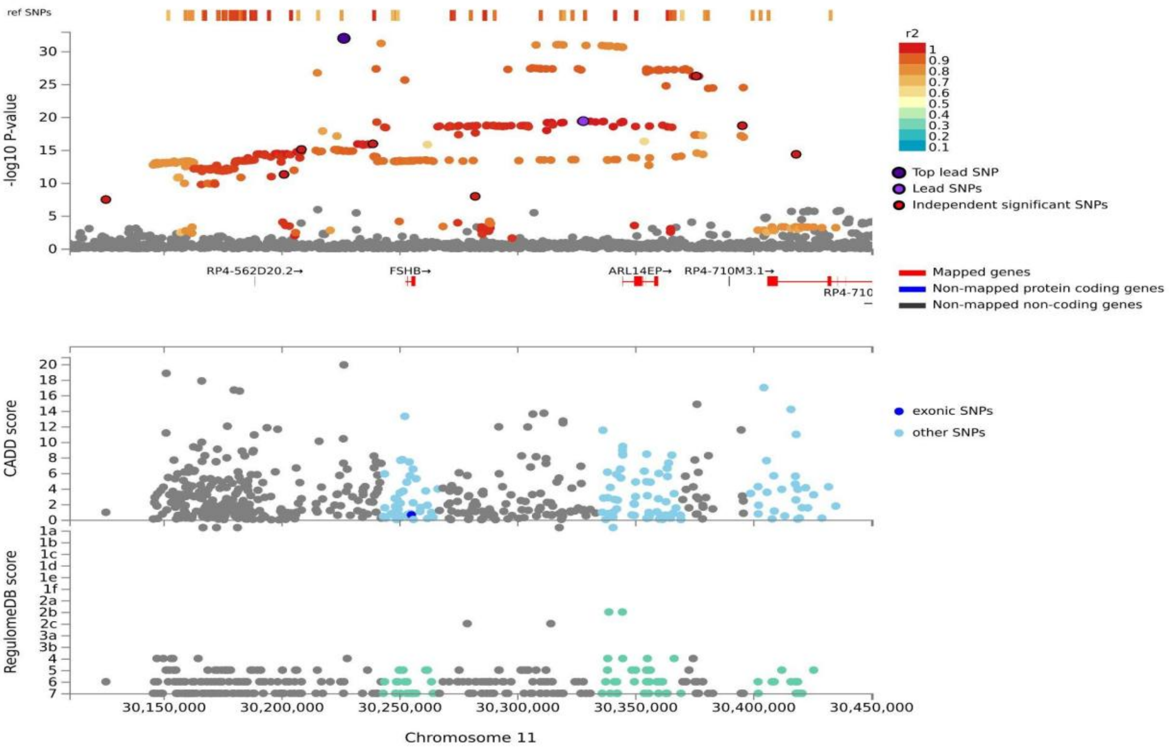


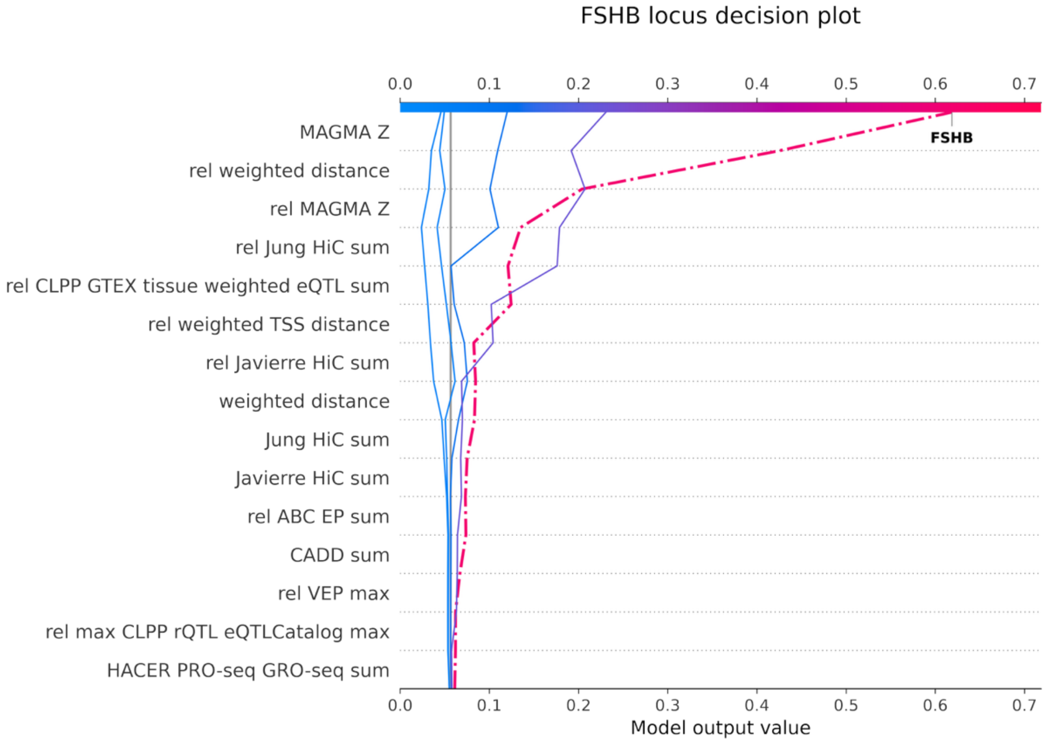

**Supplementary Fig. 6:** Locus plot and FLAMES decision plot chromosome 11 rs5030179

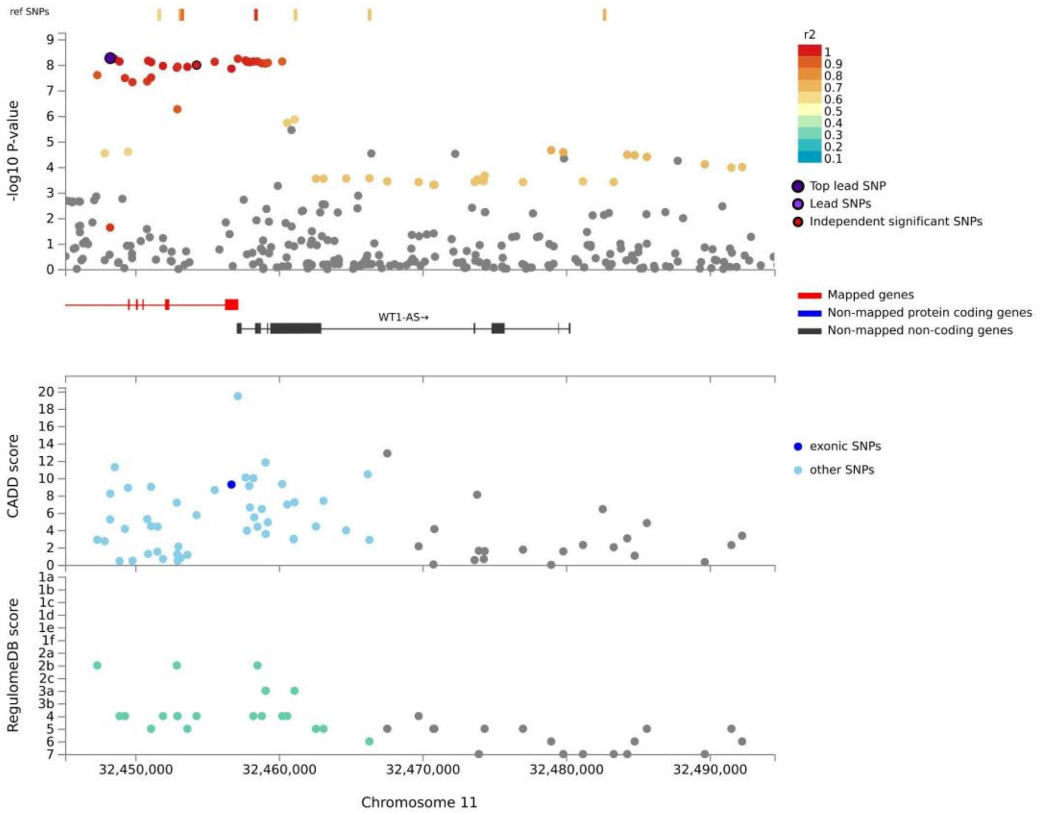


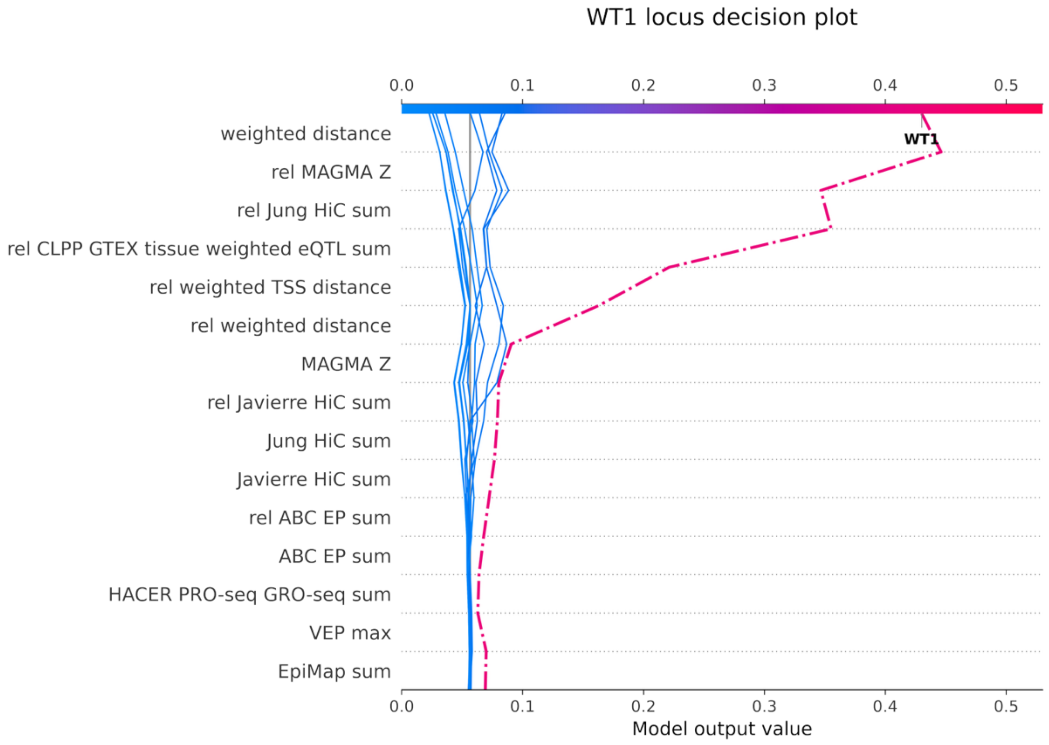

**Supplementary Fig. 7:** Results of the transcriptome-wide association study (TWAS) using FUSION[29] in the Pituitary

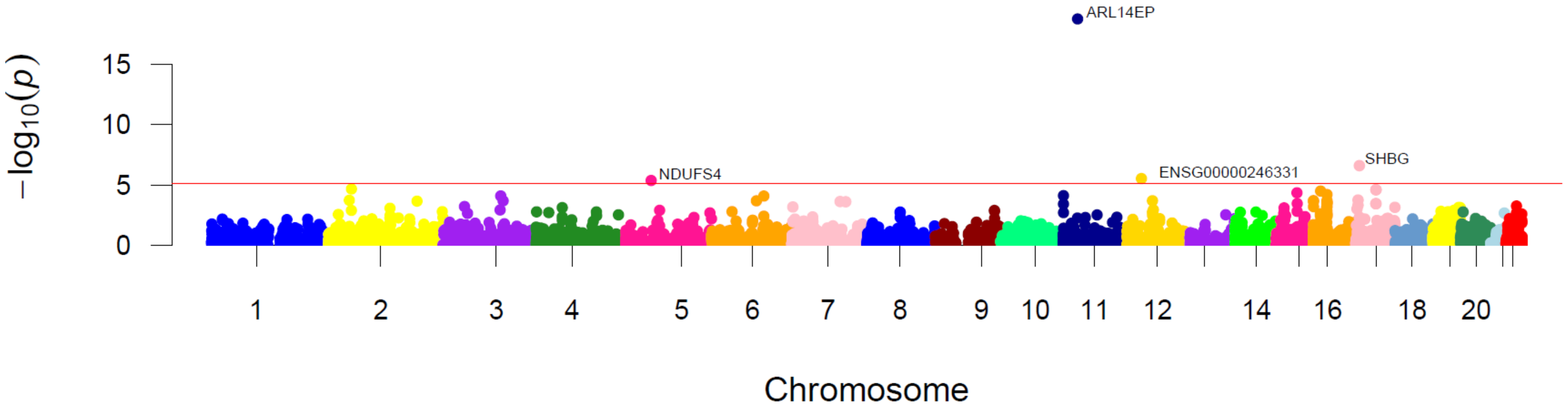

**Supplementary Fig. 8:** Results of the transcriptome-wide association study (TWAS) using FUSION[29] in the Hypothalamus

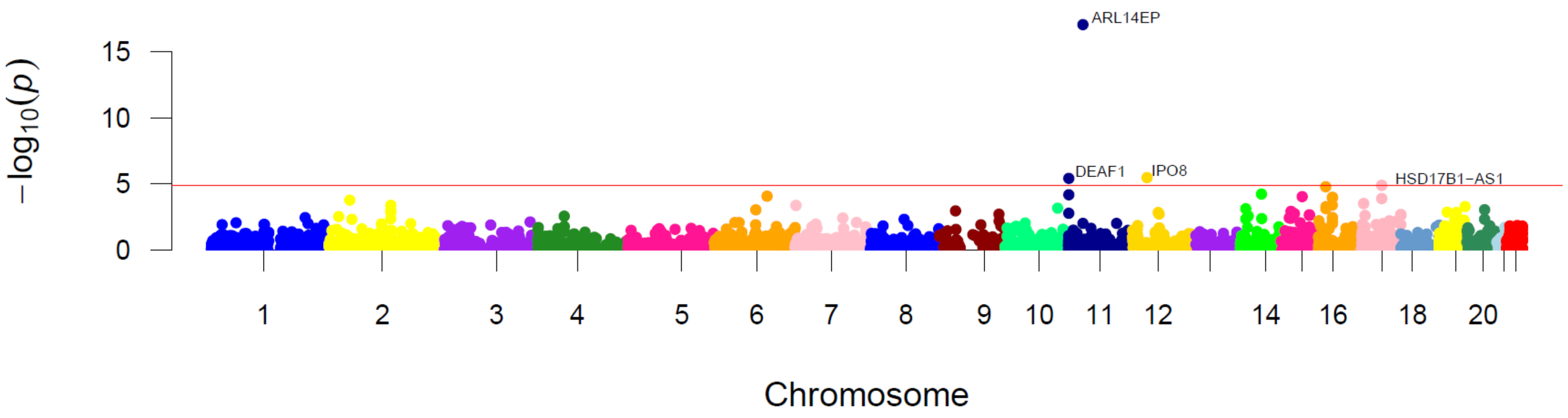

**Supplementary Fig. 9:** Results of the transcriptome-wide association study (TWAS) using FUSION[29] in Whole Blood

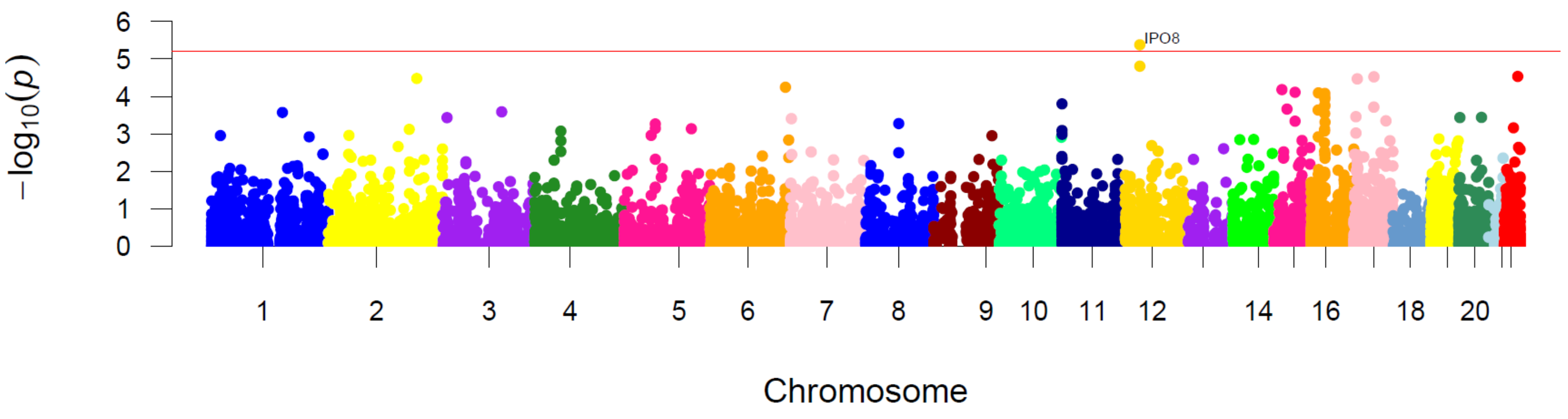

**Supplementary Fig. 10:** Results of the transcriptome-wide association study (TWAS) using FUSION[29] in the Ovary

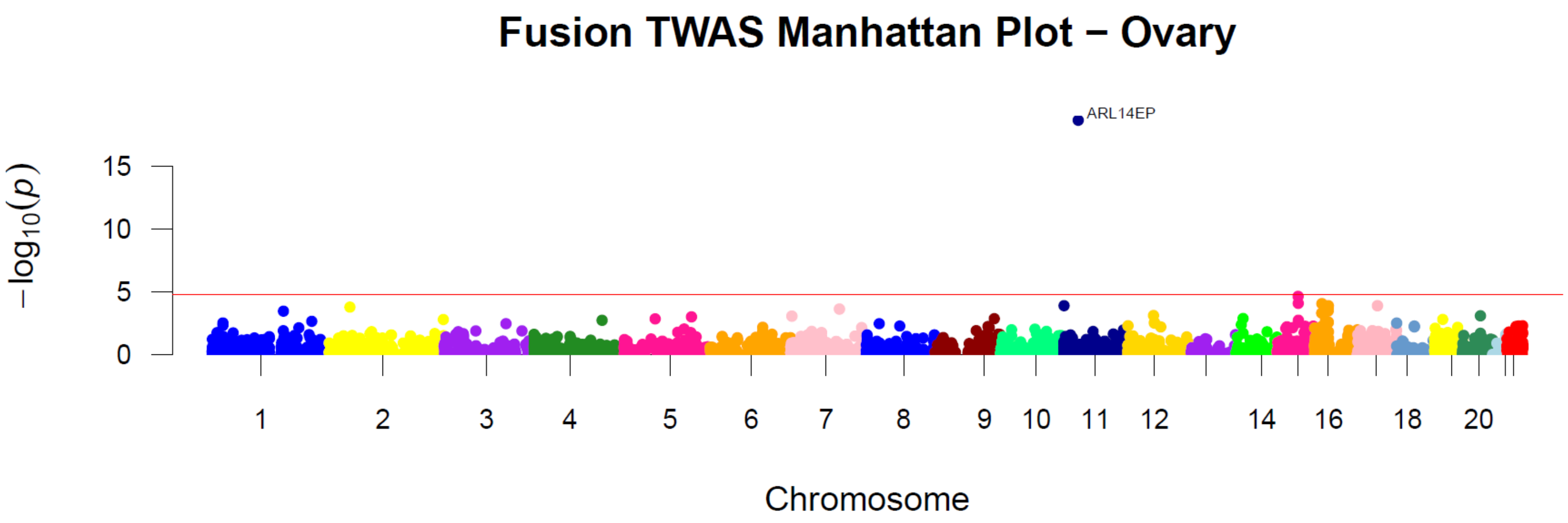

**Supplementary Fig. 11:** Results of the transcriptome-wide association study (TWAS) using FUSION[29] in the Testis

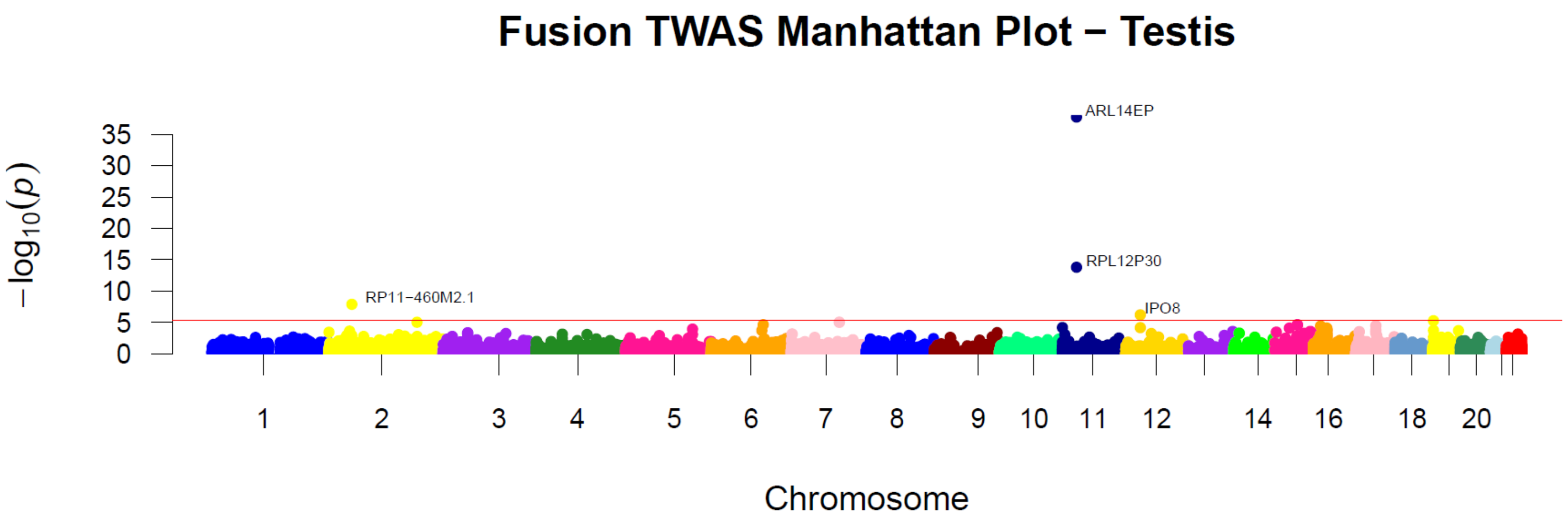

**Supplementary Fig. 12:** Results of the transcriptome-wide association study (TWAS) using FUSION[29] in the Vagina

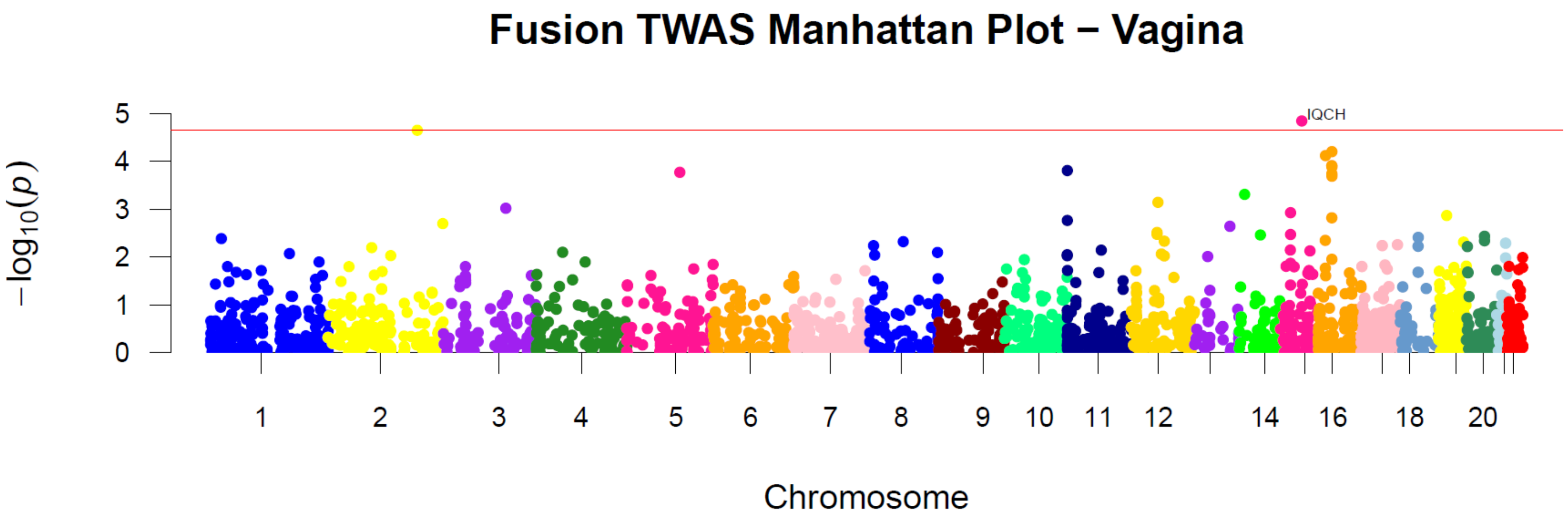

**Supplementary Fig. 13:** Results of the transcriptome-wide association study (TWAS) using FUSION[29] in Breast tissue

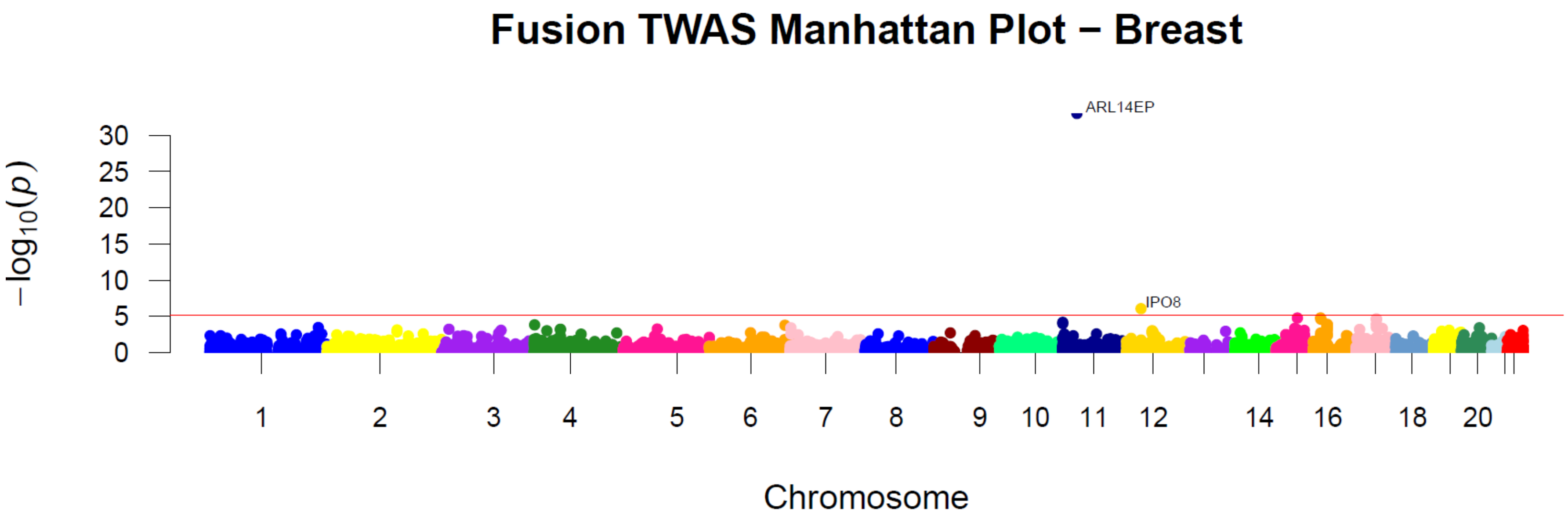

**Supplementary Fig. 14:** Results of the transcriptome-wide association study (TWAS) using FUSION[29] in the Uterus

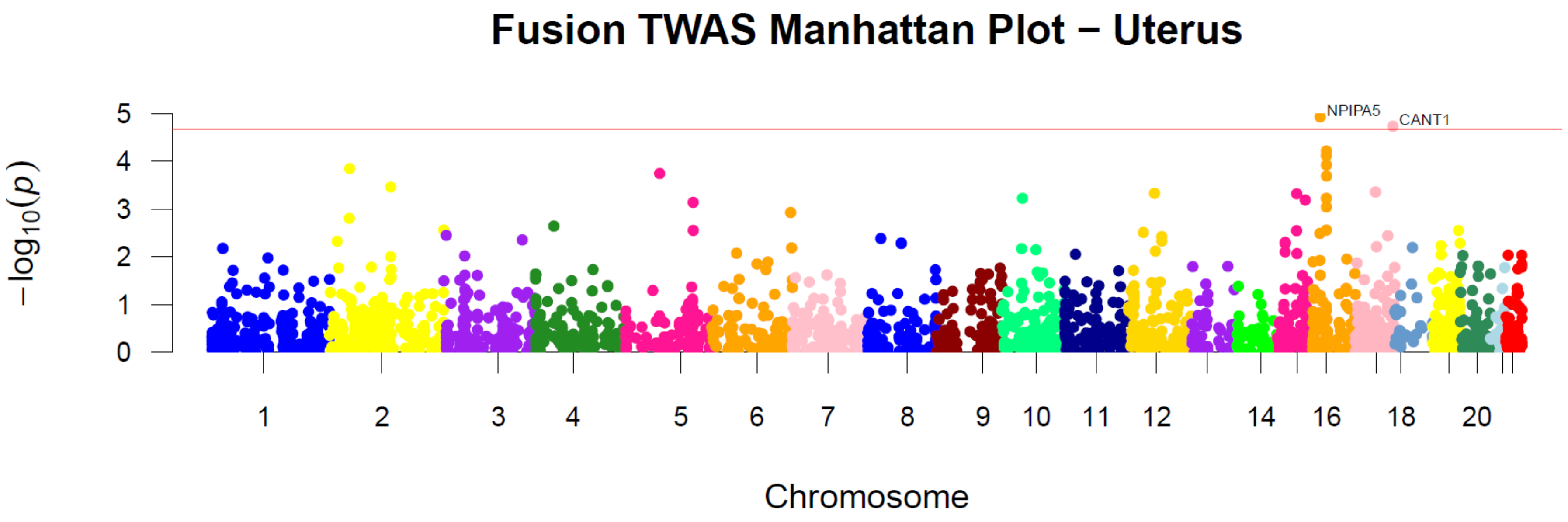

**Supplementary Fig. 15:** The burden of selection of DZT in ancient DNA with the ancient genome selection (AGES)[35] for rs11678857

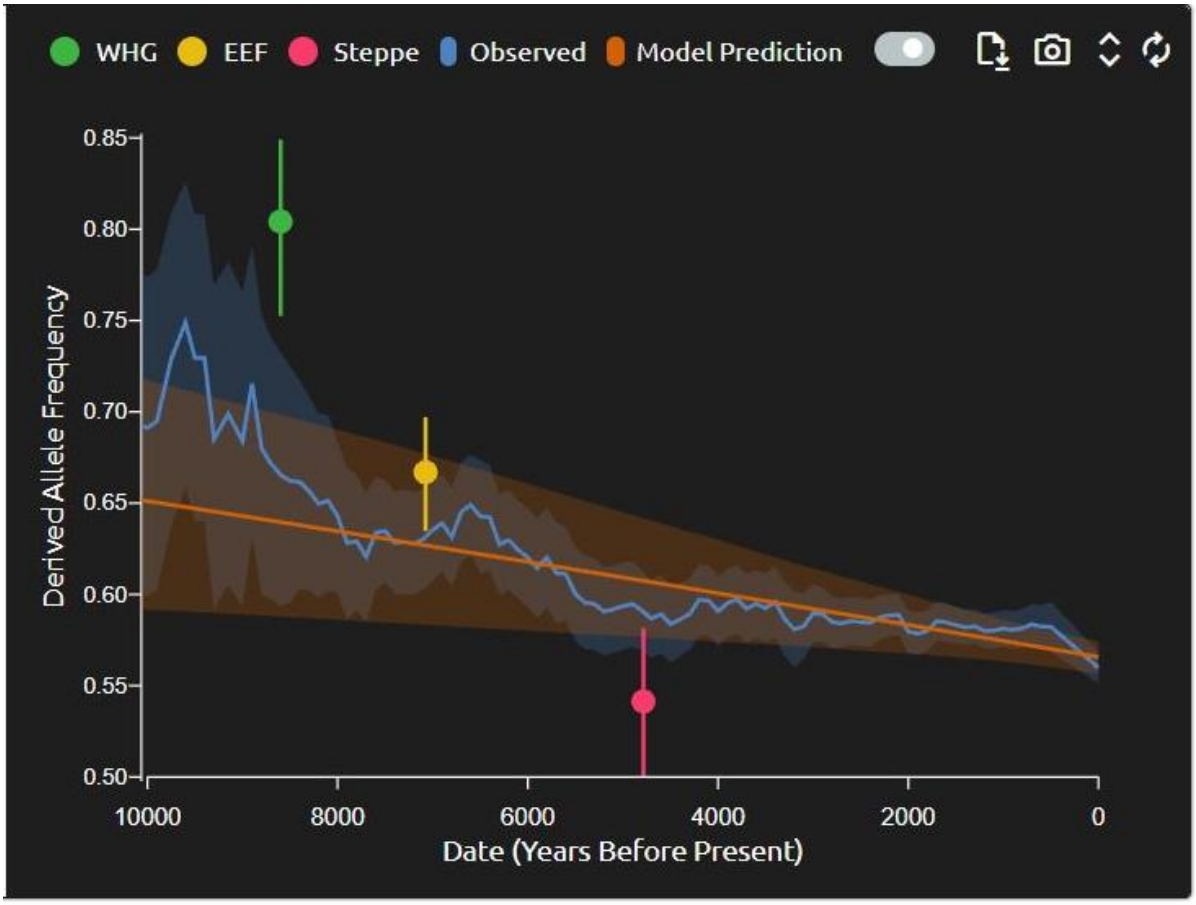

**Supplementary Fig. 16:** Cattle GWAS Q-Q plot.

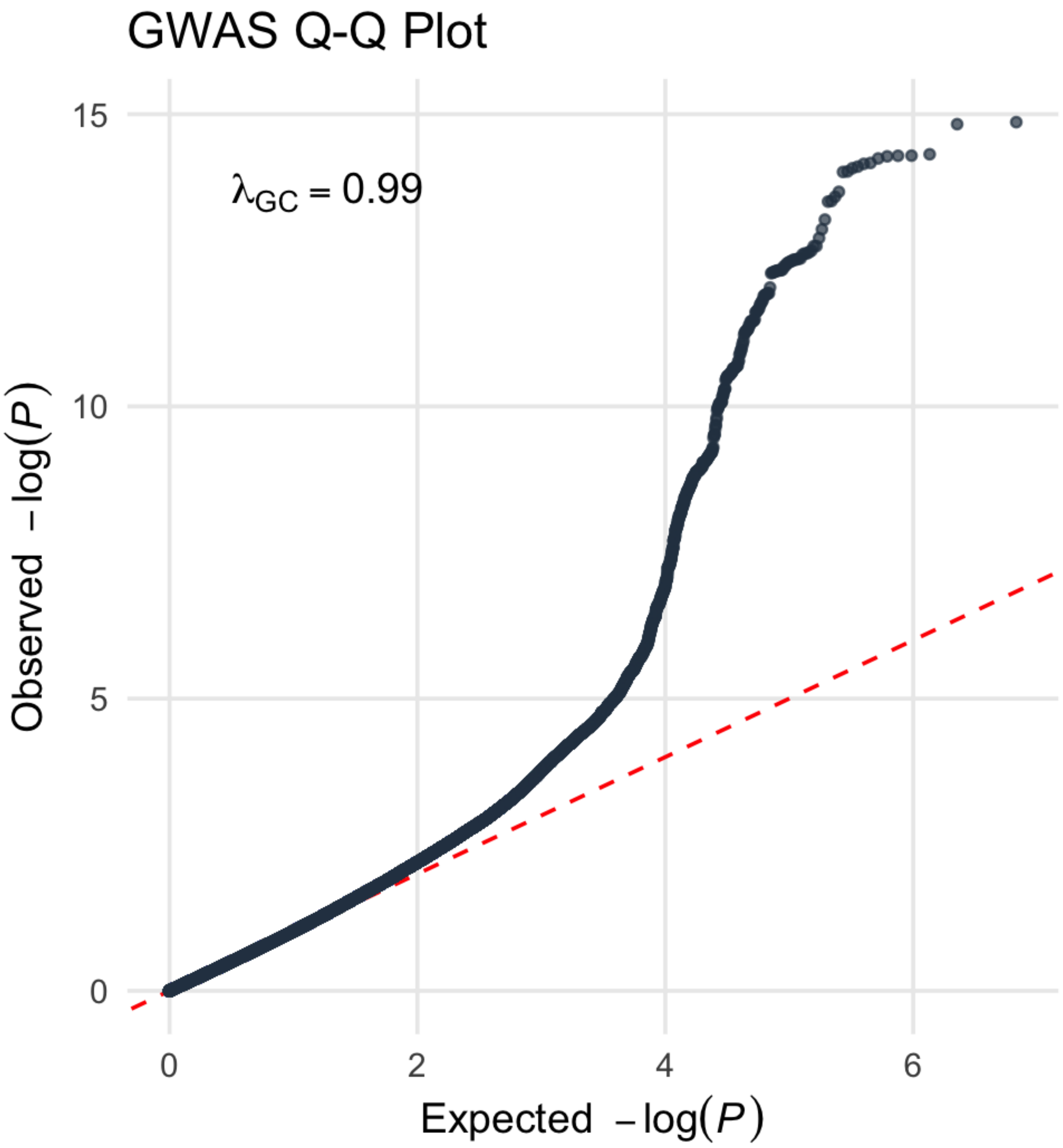
