## Supplementary Data for "Combined genome-wide association studies in humans and cattle: Cross-species genetic insights into dizygotic twinning"

**Supplementary data 1: UMCG Genetics Lifelines Initiative (UGLI) group author: LifeLines Cohort Study**

**Authors in alphabetical order**

Raul Aguirre-Gamboa (1), Roeland V. Broekema (3, 4), Patrick Deelen (1), Lude Franke (1), Jan A. Kuivenhoven (2), Esteban A. Lopera Maya (1), Ilja M. Nolte (3), Serena Sanna (1), Harold Snieder (3), Morris A. Swertz (1), Peter M. Visscher (4, 5), Judith M. Vonk (3), Cisca Wijmenga (1), Naomi Wray (5, 6), Loic Yengo (5)

*(1) Department of Genetics, University of Groningen, University Medical Center Groningen, The Netherlands*

*(2) Department of Pediatrics, University of Groningen, University Medical Center Groningen, The Netherlands*

*(3) Department of Epidemiology, University of Groningen, University Medical Center Groningen, The Netherlands*

*(4) Nuffield Department of Population Health, University of Oxford, United Kingdom*

*(5) Institute for Molecular Bioscience, The University of Queensland, Brisbane, Queensland, Australia*

*(6) Department of Psychiatry, University of Oxford, United Kingdom*

**Supplementary data 2: Standard operating procedure**

### TWINNING GWAS CONSORTIUM

### ANALYSIS PLAN

**Version:** v2 (March 2024)

**Filename:** TGC_AnalysisPlan_MA3_Complete.doc

**Attached files:**

- Surfdrive - **https://surfdrive.surf.nl/files/index.php/s/Xygww5LfZnF7NVh**
- Template_Cohort_descriptives.xlsx (Please note the three sheets)

**Deadline:** 15 July

**Prepared**: Nikki Hubers, Jouke-Jan Hottenga & Dorret Boomsma

#### BACKGROUND

The purpose of the Twinning Genetics Consortium (TGC) is to conduct meta-analyses of genome-wide association study data to identify genetic polymorphisms associated with the "Twinning" trait. Knowledge of these genes may identify key mechanisms controlling ovarian function and can provide a greater understanding of female fertility. With this SOP we are hoping to get an overview of which cohorts will join the TGC in the third Meta-analysis of Dizygotic (DZ) Twinning. We hope to increase sample size, and to include the analysis of the X-chromosome. In case of any questions please contact Nikki Hubers

##### Dizygotic twinning

Previously within TGC we were able to identify genetic locations associated with DZ twinning. The previous genome-wide association meta-analysis which included both DZ twins and mothers of DZ twins (Mbarek et al., 2024), highlighted six genes. Three, i.e., *FSHB, FSHR* and *GNRH1,* have a well-established role in female reproduction and *SMAD3* has been associated with DZ twinning in our first meta-analysis (Mbarek et al., 2016). The other two genes, *ZFPM1* and *IPO8,* have not been previously indicated. Several loci just fell short of genome-wide significance level, indicating that an increase in sample size could lead to further discoveries.

#### TRAIT DEFINITION

##### 1. Being a "mother of dizygotic twin" - MODZT

Cases: mothers of DZ twins with documented history of natural (spontaneous) DZ twinning (*i.e.,* no use of Medically Assisted Reproduction-MAR techniques)

Controls: mothers and fathers of monozygotic (MZ) twins, mothers and fathers of singletons, healthy controls, or unrelated case cohort (e.g., skin cancer cohort Australia or depression cohort NL). Please exclude participants from the controls if they have any known relative (1th and 2nd degree) that is a DZ twin or is a mother of a DZ twin. Please also check whether the genotyped cases and controls are genetically unrelated (1th and 2nd degree) e.g. with Plink IBD sharing or the King genetic relationship software.

##### 2. Being a "dizygotic twin" - DZT

This study design is only suitable when just the DZ twins are genotyped and not their mothers (i.e., twin data are only included if the mothers were not genotyped).

Cases: spontaneous DZ twin (*i.e.,* mothers did not use of Medically Assisted Reproduction-MAR techniques)

Controls: non-DZ twins: MZ twins, mothers and fathers of MZ twins, singletons, mothers and fathers of singletons, healthy controls, or unrelated case cohort (e.g., skin cancer cohort Australia or depression cohort NL). Please exclude participants from the controls if any known relative (1th and 2nd degree) is known to be a DZ twin, or is a mother of a DZ twin, or any genetically 1st and 2nd degree related people to all cases.

**In case your cohort has both MODZT and DZT cases we ask you to first focus on the MODZT cases. If you have enough independent cases of DZT (e.g. no mothers genotype present) and independent controls, please include both analyses separately.**

**Important notes:**

- The minimum number of cases in the analyses should be N=100, preferably with at least the same number (or more) of controls.

- Zygosity of the twins should be checked by preferably genotypic data, or if unavailable, blood group or extensive survey information.

- We ask for strict selection of the cases as the inclusion of mothers who conceived their twins through MARs diminishes the signal drastically (see Mbarek et al. 2016). Exclusion criteria can be based on medical records, targeted questions on the mode of conception, or the birth year of the twins (e.g., fertility treatments were started in the Netherlands in 1970, so mothers of twins born before 1970 can be included). Please, indicate how MAR use was determined in the Study descriptives file. Still, we ask you to (if you are collecting data) keep collecting MAR participants. In further work we would like to look into the genetics of MAR twin mothers.

- Large epidemiologic studies have found no etiological connection between DZ and MZ twinning. Thus, (parents of) MZ twins form useful controls and we shall use both mothers and fathers where available.

- For controls, exclude all participants when there are DZ twins, mothers of DZ twins and children of DZ twins in the family / pedigree (if possible).

#### EXCLUSION CRITERIA

- Unclear zygosity
- Unclear MAR status

#### COVARIATES

- 10 Principal Components calculated with projection to the 1000 genomes as described at https://www.biostars.org/p/335605/. We can also provide a less computational intensive code with the GWAS SOP upon request.
- Micro-array or other technical covariates
- Study specific covariates

#### GENOTYPES & IMPUTATION

In case you are still planning to do your imputation we would recommend you to use the HRC reference panel using build 37 and follow the instruction of the HRC imputation server (Getting Started - Michigan Imputation Server). If you are unable to upload your genotypes due to privacy restrictions please follow the **same** data cleaning preparation processes as imputation server does (Data Preparation - Michigan Imputation Server) and then use the latest available in house reference panels preferably build 37, e.g. look at (IGSR | data collections (internationalgenome.org / Haplotype Reference Consortium Release 1.1 - EGA European Genome-Phenome Archive (ega-archive.org)). We recommend imputation software packages such as IMPUTE5, EAGLE + MACH4, or Beagle.

If you are planning on performing the imputation, please keep the following QC checks in mind:

Remove samples that have:

- low overall sample call-rate (<95%)
- excess autosomal heterozygosity (Plink |Fhet<0.1| or please specify)
- Duplicate samples
- mismatch between self-reported and genetic sex (excessive X-chromosome heterozygosity in individuals classified as males)
- XXY genotype and other aneuploidies.
- Please check the relatedness of sample using the King software (KING: Relationship Inference Software (kingrelatedness.com)).

Remove SNPs that have:

- SNP Call rate <95% - 98%
- HWE p < 1e-6 up until 1e-4.
- Known to have evidence of poor clustering based on genotyping software.
- Palindromic A/T G/C SNPs with allele frequencies between 0.30-0.70.

After these checks you can start with Data Preparation - Michigan Imputation Server. This will align the sample SNP data with the reference panel and that is very important.

In case of questions concerning imputation and your current data being sufficient, please contact Jouke-Jan Hottenga. We can provide aid and scripts for imputation upon request.

After imputation, please check the imputation logs. Important things to check are mis-alignment of strand between the reference and input data (especially with palindromic SNPs) and a comparison of the number of SNPs that went into the imputation and the number of SNPs that were actually used during the imputation (e.g. Beagle tends to not use the SNPs where reference and alternate allele are swapped. Use plink –keep-allele-order or assign either ref. or alt. allele as allele 1 or 2 to fix this.). Furthermore, it is wise to make a plot comparing the imputed allele frequencies of the sample to the reference data. This should be cigar shaped, with zero - or very few outliers. Any Z or X shaped plots means that SNP alignment steps failed to work and that will require a new imputation.

#### MODELS USED TO TEST ASSOCIATION

A GEE Logistic regression model will be performed on all samples adjusted for appropriate covariates. After consideration, we have opted to perform the analyses in fastGWA-BB from the GCTA software package (https://yanglab.westlake.edu.cn/software/gcta/#Overview). Please note that this tool requires a premade generic relatedness matrix (GRM).

We provide full explementary scripts for making the GRM and performing the GWAS, which can be found in the Twin_pipeline_MA3 folder. To access the pipeline please download all tools and scripts to a linux environment and don't forget to use the command chmod +x to make all the files and tools executable. For any questions regarding the pipeline please contact Nikki Hubers

#### Models:

**GWAS DZ twinning**

1. **MOTHDZT *vs* controls**

Logit (MOTHDZT vs controls)= SNP + PC's + Study specific covariates + GRM

2. **DZT vs controls**

Logit (DZT vs controls)= SNP + PC's + Study specific covariates + GRM

**Notes:**

- **Run analyses on all 22 autosomes and the X chromosome. For chromosome X, please run an analysis in females only.**
- Additional and more detailed instructions can be found in the comments of the .sh script files.

**Needed for upload:**

The 23 MOTHDZT fastgwa output files.

The 23 DZT fastgwa output files.

All logfiles of the GWAS analysis.

The cohort descriptes Excel file, template: Template_Cohort_descriptions.xlsx

**DELIVERY**

We will ask you to deliver your complete GWAS results by: **15 July 2024.** A completed Template_Cohort_descriptives.xlsx file and a gziped version of the **yymmdd**_TWIN_AllCHR_**COHORTNAME**.dat.gz file can be uploaded in the surfdrive**:** **https://surfdrive.surf.nl/files/index.php/s/Xygww5LfZnF7NVh.** The password for the surfdrive was provided in another email. In case of any questions please contact Nikki Hubers.
